# Global gram-negative diabetic foot infection prevalence and its associations with climate: a systematic review and meta-analysis

**DOI:** 10.1101/2025.02.15.25322334

**Authors:** Marcos C. Schechter, Ben Rabin, Ellen Martinson, Mia S. White, Baffour Otchere, J. Raymond, Jillian Dunbar, Meg McAloon, Priyanka A. Bhanushali, Kyra L. Urquhart-Foster, Maya Fayfman, Mohammed K. Ali, Eric Senneville, Rodrigo M. Carrillo-Larco, Sophie Lockwood

## Abstract

**Background:** Based on limited data, the International Working Group on the Diabetic Foot guidelines recommend antibiotics with anti-gram-negative activity, including against P. aeruginosa, for patients with moderate-to-severe diabetic foot infections (DFIs) who reside in warm climates and state that P. aeruginosa DFIs are more common in North Africa/Asia vs Western Europe/North America.

**Methods:** We performed a systematic review and meta-analysis of studies of any design and language published between January 1, 2010, and December 15, 2020, that reported the proportion of participants with DFIs and microbiological tests that were positive or negative for *Pseudomonas aeruginosa* and/or gram-negatives of any species. We calculated the pooled prevalence of *P. aeruginosa* DFIs (psaDFIs) and overall gram-negative DFIs (gnDFIs) defined as the proportion of DFIs presenting to healthcare facilities with microbiologically-confirmed DFIs using random effects models. This review is registered with PROSPERO (CRD42022279019).

**Findings:** We identified 246 studies (36,788 participants) reporting on psaDFI prevalence and 55 studies (6,079 participants) reporting on gnDFI prevalence. The psaDFI pooled prevalence in tropical, subtropical, and temperate zones was 19% (95%CI 17-22%), 14% (95%CI 12-17%), and 14% (95%CI 11-17%), respectively. The gnDFI pooled prevalence in tropical, subtropical, and temperate zones was 68% (95%CI 61-74%), 54% (95%CI 45-63%), and 43% (95%CI 35-52%), respectively. The pooled prevalence of psaDFIs and gnDFIs was higher in studies conducted in Asia/North Africa [psaDFI 19% (95%CI 17-21%) and gnDFI 57% (95%CI 51-64%)] compared to studies conducted in North America/Western Europe [psaDFI 13% (95%CI 10-17%) and gnDFI 42% (95%CI 32-52%)]. However, the pooled prevalences of these infections were similar in temperate zones of these regions. Few studies reported data on key risk factors for psaDFIs and gnDFIs, including DFI severity and prior antibiotic exposure, and there was substantial between-study heterogeneity within climate zones and regions.

**Interpretation:** psaDFIs and gnDFIs were more prevalent in tropical zones compared to temperate zones, but not subtropical zones. Climate zones better discriminate DFI microbiology compared to the guideline-proposed regional differences. Available studies lack necessary data to separate the effects of climate from other risk factors for psaDFIs and gnDFIs.

**Funding:** None

**RESEARCH IN CONTEXT:** *Evidence before this study:* Diabetic foot ulcers are a leading cause of preventable limb loss globally and diabetic foot infections (DFIs) are generally the terminal step before an amputation. Whether or not to use antibiotics with anti-gram-negative activity, particularly anti-*Pseudomonas aeruginosa*, is a key DFI management decision. The International Working Group on the Diabetic Foot is an international multidisciplinary organization that issues diabetic foot-related guidelines adopted globally. The latest guidance issued in 2023 states that the decision to use anti-gram-negative and anti-pseudomonal agents should be based on infection severity, recent antibiotic exposure, prior culture data, and the climate and location where the patient resides (table). The climate- and location-related recommendations are based on limited data. We searched PubMed, EMBASE, Web of Science, and World Health Organization databases on December 15, 2020, to identify articles published between January 1, 2010, and December 15, 2020, and repeated the PubMed search on December 30, 2024. Two prior systematic reviews and meta-analysis found higher *P. aeruginosa* DFI (psaDFI) rates in Asia and Africa compared to the Americas and Europe. We found no prior systematic reviews reporting on the prevalence of psaDFIs and overall gram-negative DFIs (gnDFI) globally stratified by climate zones.

*Added value of this study:* From 246 studies (36,788 participants) reporting on psaDFI prevalence and 55 studies (6,079 participants) reporting on gnDFI prevalence, psaDFIs and gnDFIs were more prevalent in tropical settings compared to temperate zones, but there were no substantial differences in the prevalence of these infections between tropical and subtropical, and subtropical and temperate zones. Additionally, we found that climate zones better discriminate the prevalence of these infections compared to the guideline-proposed regional differences. Importantly, studies generally lacked key data needed to understand if the differences in prevalence were related to characteristics other than climate (DFI severity, antibiotic exposure) and there was substantial between-study heterogeneity within climate zones and regions that could not be explained by meta-regression models that included climate, region, or inpatient *vs* outpatient recruitment (a proxy for DFI severity).

*Implications of all the available evidence:* Well-designed geographically representative cohort studies that contain patient-level data regarding the contribution of location and clinical factors on DFI microbiological profile and treatment outcomes are needed to better guide antimicrobial therapy. Absent further evidence, the future guidelines could state psaDFIs and gnDFIs are more common in tropical zones. Table:
International Working Group on the Diabetic Foot/Infectious Diseases Society of America 2023 recommendations regarding use of anti-gram-negative, including anti-pseudomonal, antibiotics.

|  |
| --- |
| Target aerobic gram-positive pathogens only (beta-haemolytic streptococci and Staphylococcus aureus including methicillin-resistant strains if indicated) for people with a mild DFI, who have not recently received antibiotic therapy, and who reside in North America or Western Europe |
| Do not empirically target antibiotic therapy against Pseudomonas aeruginosa in cases of DFI in temperate climates, but use empirical treatment of P. aeruginosa if it has been isolated from cultures of the affected site within the previous few weeks, in a person with moderate or severe infection who resides in Asia or North Africa. (Note: the guidelines suggest using anti-pseudomonal agents for patients with moderate or severe DFIs with macerated ulcers or who live in warm climates). |

## INTRODUCTION

Diabetes-related lower-extremity complications are a leading cause of disability globally^1^. In 2016, 18.6 million people had a diabetic foot ulcer (DFU) and 6.8 million people were living with a diabetes-related amputation^1^. Unfortunately, DFUs and amputation burdens are expected to increase as diabetes surges globally^2^. The pathway to diabetes-related amputation is complex, and involves an interplay of biomechanical factors, neuropathy, and peripheral artery disease (PAD), but diabetic foot infections (DFIs) are generally the terminal step before limb loss^3,4^. Antibiotic therapy is a cornerstone of limb salvage following a DFI, but antibiotic choice for DFIs is complex because they are generally polymicrobial and obtaining adequate specimens for microbiological tests is challenging^5,6^.

Whether or not to use antibiotics with anti-gram-negative activity, particularly anti-*Pseudomonas aeruginosa*, is a key DFI management decision^5^. Current international guidelines state the decision to use anti-gram-negative and anti-pseudomonal agents should be based on infection severity, recent antibiotic exposure, prior microbiological data, and the climate and location where the patient resides. Increased infection severity and prior antibiotic exposure are associated with increased prevalence of gram-negative (including *P. aeruginosa*) DFIs, while prior isolation of gram-negative or *P. aeruginosa* are associated with increased odds of gram-negative or *P. aeruginosa* DFIs. Regarding climate, the guidelines recommend anti-gram-negative antibiotics with anti-pseudomonal activity for moderate and severe DFIs for patients who live in warm climates. Regarding location, the guidelines state that *P. aeruginosa* DFIs are more common in North Africa and Asia compared to Western Europe and North America. However, these climate- and location-related recommendations are based on non-systematic comparisons between cohorts. For example, the guidelines cite studies conducted in the United States (US)^7^ and Turkey^8^ that enrolled participants with moderate-to-severe DFIs where the gram-negative and *P. aeruginosa* rates were lower in the US compared to Turkey (14% *vs* 60.2% for gram negative and 2.5% *vs* 12.4% for *P. aeruginosa*).

The afore-mentioned non-systematic comparisons are limited because they do not incorporate the full body of the literature and do not account for differences between cohorts, such as differences in DFI severity or prior antibiotic exposure. To our knowledge, there are no published systematic reviews and meta-analysis investigating the associations between climate and DFI microbiology; here, we aim to fill this knowledge gap.

## METHODS

### Search strategy, Study Selection, and Data Abstraction

We searched PubMed, EMBASE, Web of Science, and World Health Organization databases (AIM, IMSEAR, IMEMR, LILACS, and WPRIM) on December 15, 2020, to identify articles published between January 1, 2010, and December 15, 2020. We used the search terms “diabetic foot” AND “infection” or “soft tissue infection” or “osteomyelitis” and related terms (supplemental file). There were no restrictions placed on publication language.

Studies of any design that reported the proportion of participants of any age with a DFI who had a lower-extremity microbiological test negative or positive for *Pseudomonas aeruginosa* and/or a gram-negative bacillus of any species were eligible for inclusion. Microbiological tests could be from soft tissue and/or bone and culture- and/or molecular-based. We excluded studies that had <5 participants and conference abstracts. Our primary objective was to investigate the prevalence across climates (defined as the proportion of DFIs presenting to healthcare facilities with microbiologically-confirmed *P. aeruginosa* and/or gram-negative infection). We excluded studies that (1) reported multiple specimens per participant, (2) included participants from >1 climate zone and did not stratify results by climate zone, and (3) did not report the number participants that tested negative for *P. aeruginosa* and/or a gram-negative infection. If multiple publications from the same center met inclusion criteria, we reviewed the recruitment dates to determine if there could be overlap between participants and queried the study. Where overlap was confirmed or could not be excluded, we retained the study with the largest study population.

Titles, abstracts, and full texts were screened by two reviewers independently, and discrepancies were resolved through discussion between the reviewers and/or by a senior investigator (MCS). We extracted all data into standardized forms using double data entry. We used Google Translate to review studies whenever assigned reviewers could not read the original publication language.

### Outcome and Exposure Abstraction and Definitions

We followed the PICO format. The population was participants with a DFI (soft tissue and/or osteomyelitis), and the intervention was a microbiological test (culture and/or molecular). The main comparison was the climate where the study was done (tropical *vs* subtropical *vs* temperate)^9^, and the secondary comparison was the region where the study was done (North Africa/Asia *vs* North America/Western Europe). Finally, the outcomes were culture-confirmed *P. aeruginosa* DFIs (psaDFIs) and gram negative DFIs of any species, including *P. aeruginosa* (gnDFIs).

For each included study, we abstracted the proportion of participants with a DFI who had a positive microbiological test by culture and/or molecular methods from a foot sample (however defined by the investigators) for *P. aeruginosa* and/or gram-negative bacilli of any species. Most studies reported data for participants with positive and negative microbiological tests in aggregate. Whenever available, we used the number of patients with a positive microbiological test as the meta-analysis denominator. If we were unable to determine the prevalence of DFI involving gram-negative bacteria, we attempted to contact the original study authors to obtain this data.

If a study reported microbiological data from both cultures and molecular tests, we used the culture results which are favored by current guidelines^5^. DFIs occur in a spectrum that range from superficial soft tissue to deep soft tissue to bone infection (osteomyelitis). There is limited agreement between wound swabs and tissue samples among persons with soft tissue DFIs, and between soft tissue and bone samples among persons with diabetic foot osteomyelitis^6,10^. If a study reported multiple sample sources, we used the deepest sample reported for our analysis.

For our primary analysis, we classified climate based on latitude boundaries and classified them as tropical (23.4 to -23.4 degrees latitude), subtropical (23.4 to 35 degrees and -23.4 to -35 degrees latitude), and temperate (between 35 to 66.5 degrees and -35 to -66.5 degrees latitude)^9^. In a secondary analysis, we compared the prevalence of psaDFIs and gnDFIs between North Africa/Asia and North America/Western Europe based on WHO definitions^11^ . The latitude boundaries data abstraction and definitions North Africa/Asia and North America/Western Europe are in supplemental tables 1 and 2.

### Study Risk of Bias Quality Assessment

Since we included reports of varying study designs, we developed a set of questions to assess risk of bias and quality tailored to our study aims. We abstracted data regarding (1) demographics, (2) participant recruitment, (3) antibiotic exposure prior to microbiological testing, and (4) DFI severity, and assigned study quality indicators for each of these 4 domains. For each of these four criteria, we defined a 3-point scale based on data completeness which are summarized in supplemental table 3.

The demographic and participant’ recruitment indicators were selected to investigate if the study participants were likely to reflect the general population with DFIs (i.e., risk of selection bias). For the demographic study quality assessment, studies were classified as (1) “complete data” if (a) the proportion of males and females *and* (b) the study mean or median age was reported, (2) “incomplete data” if only one of these data were reported, and (3) “no data” if none of these data were reported. The best metric was “complete data”.

The participant recruitment study quality assessment was defined as (1) “consecutive” if the study population consisted of all consecutive eligible patients with a DFI that met the original study inclusion criteria, (2) “random” if the study population consisted of a random subset of patients with a DFI, and (3) “convenience or missing” if the study population was selected by convenience sampling or by unclear methods. The best metric was “consecutive”.

The antibiotic exposure study quality assessment was defined as “complete data” if (a) the proportion of the study population exposed to antibiotics within a defined period prior to microbiological testing was reported *and* (b) the microbiological results were stratified by prior antibiotic exposure. The DFI severity quality assessment was defined as “complete data” if (a) the proportion of patients with severe, moderate, or mild infection was reported *and* (b) the microbiological results were stratified by DFI severity^5^. DFI severity was based on the IDSA/IWGDF, Wagner, and/or University of Texas classification systems(described in the supplemental table 4). The antibiotic exposure and DFI severity quality assessments were defined as “incomplete data” if the proportion of the study population exposed to antibiotics within a defined period, or the proportion with severe, moderate, or mild infection was reported but microbiological results were not stratified by these characteristics. Finally, the quality assessments for these indicators -prior antibiotic exposure and DFI severity-were defined as “no data” if the proportion of the study population with these characteristics was not reported. The best metric for these indicators was “complete data”.

### Other Variables Abstracted

As few studies reported DFI severity, we recorded where participants were recruited, including exclusively in outpatient clinics, exclusively in inpatient or emergency departments, or in both outpatient and inpatient facilities as DFI severity is likely higher among inpatient and emergency department participants compared to outpatient participants.

### Statistical Analysis

The pooled prevalence of psaDFIs and gnDFIs was calculated using random effects generalized linear mixed models on log-transformed proportions and represented by forest plots with 95% confidence intervals stratified by climate zones and regions. Between-study heterogeneity was assessed using the I^2^ statistic and the Cochrane Q statistic was calculated to test for heterogeneity. Publication bias was assessed using visual inspection of funnel plots and the Egger’s test.

We conducted a meta-regression to evaluate the role (1) climate, (2) region, (3) study setting (exclusively outpatient *vs* not exclusively outpatient), and (4) study recruitment mid-point in psaDFI and gnDFI prevalence heterogeneity. The study setting was included as a proxy for DFI severity due to the high rate of missingness for this variable among included studies. The meta-regression models included the following variables (1) climate zone or region, (2) climate zone or region and study setting (3) study recruitment mid-point. We did subgroup analysis restricted to studies from Western Europe/North America and North Africa/Asia. The amount of between-study heterogeneity accounted by these models was assessed by the R^2^ statistic.

All analyses were conducted using R version 4.3.2, including the meta^12^ and metafor^13^ packages for meta-analysis and meta-regression, respectively. We followed the PRISMA guidelines for systematic reviews and meta-analysis^14^ (checklist in the supplemental file) and registered the review on the PROSPERO database (CRD42022279019). The data and R code are in the supplemental files.

## RESULTS

### Overall Included Studies Characteristics

We screened 4,101 unique studies of which 263 met inclusion criteria (Figure 1). Included studies were published in 13 languages and represented data from 51 countries (supplemental table 5). Maps depicting included study locations are shown in Figure 2. Among the 263 studies included, 84 studies (n=11,329 participants) were from a tropical zone, 77 studies (n=10,510 participants) were from a subtropical zone, and 102 studies (n=16,541 participants) were from a temperate zone. Regarding study region, 51 studies (n=9710 participants) were from Western Europe or North America and 177 studies (n=24567 participants) were from Asia or North Africa. Among the studies from Western Europe or North America, 9 studies (n=1661 participants) were from a subtropical zone and 42 studies (n=8049 participants) were from a temperate zone. Among the studies from Asia or North America, 68 studies (n=9410 participants) were from a tropical zone, 62 studies (n=8321 participants) were from a subtropical zone, and 47 studies (n=6836 participants) were from a temperate zone.

**Figure 1:**
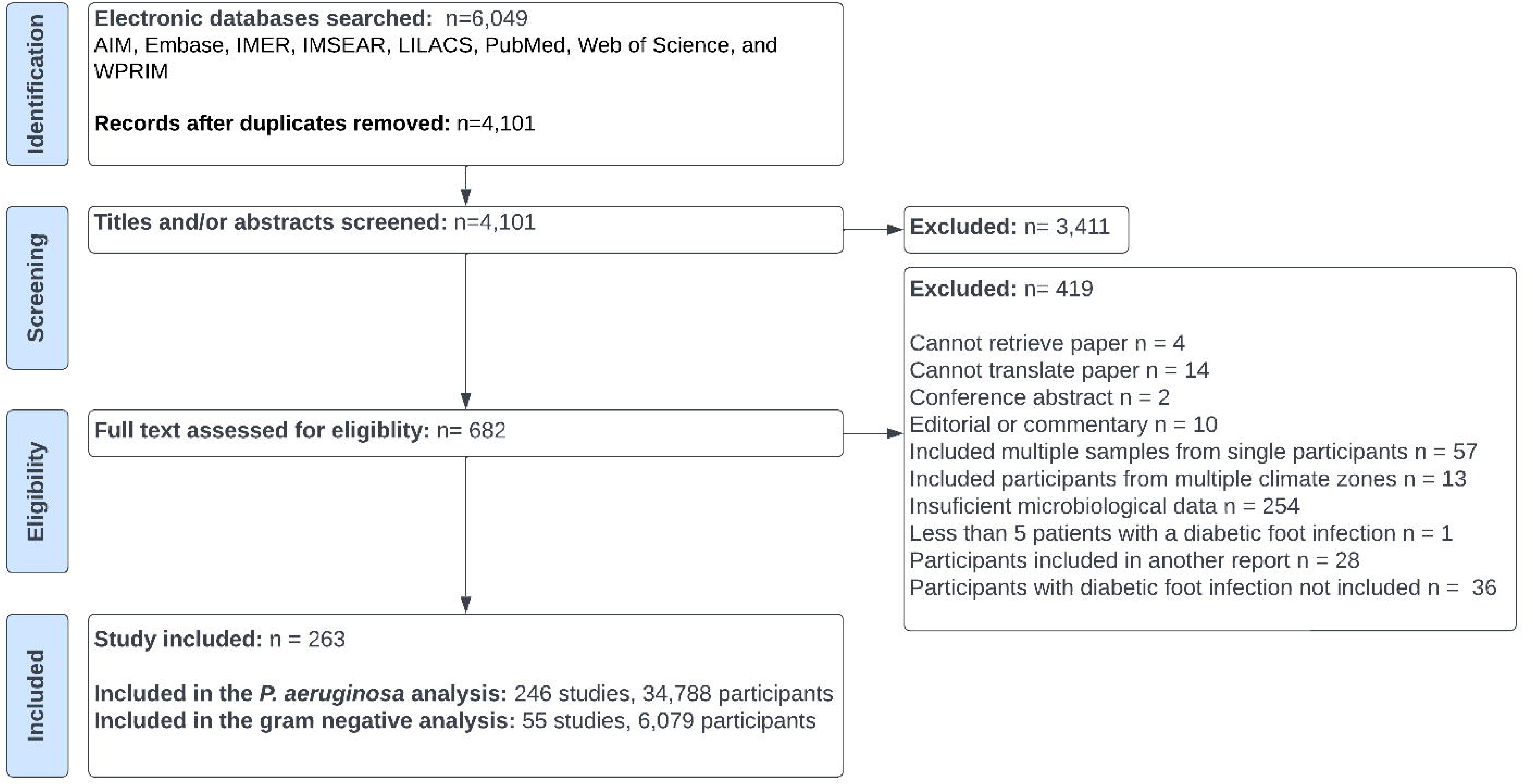
PRISMA flow diagram.

**Figure 2:**
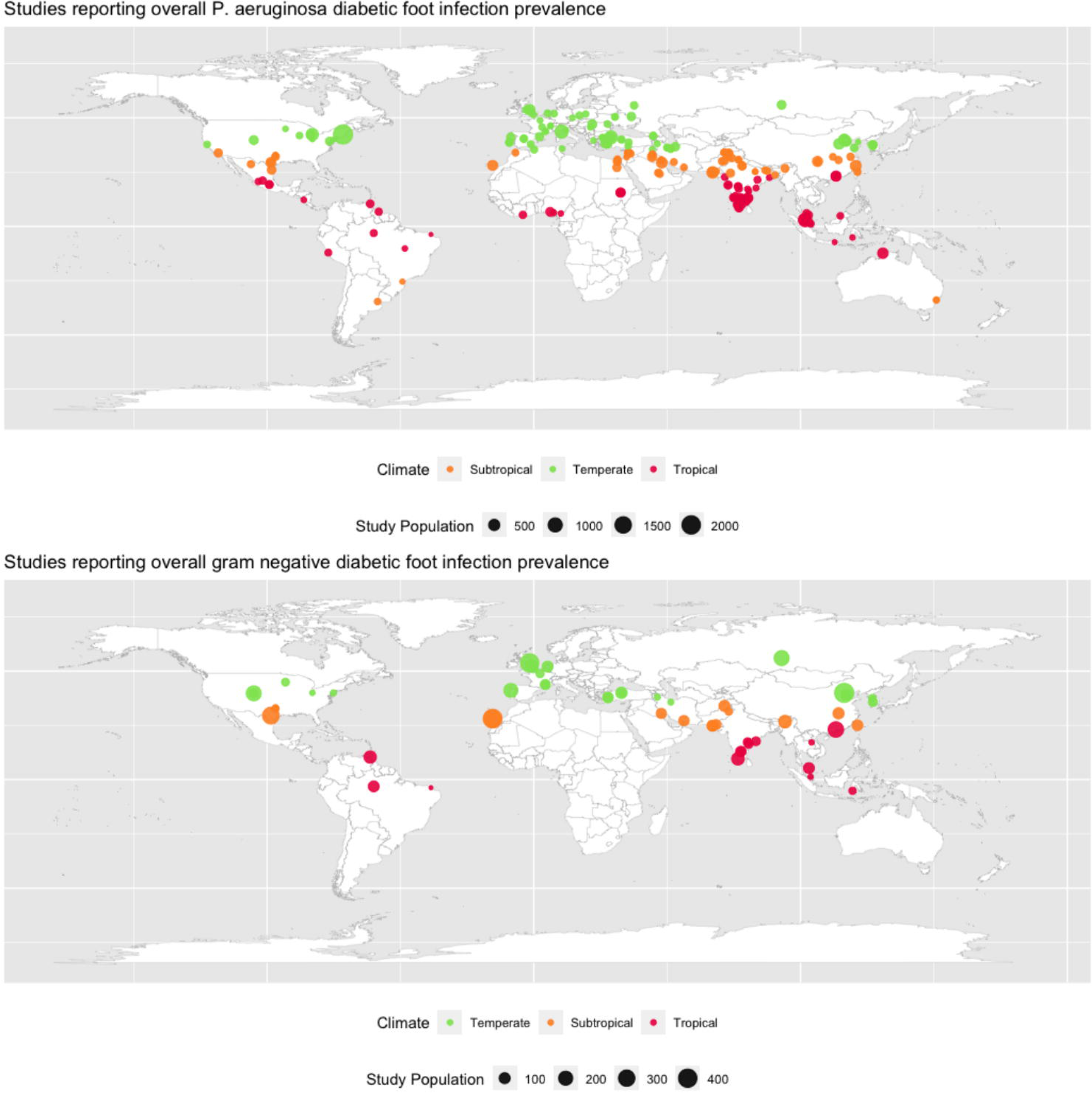
Location of the included studies.

Two-hundred and forty-six studies that included a total of 34,788 participants were included in the *P. aeruginosa* meta-analysis and 55 studies that included a total of 6,079 participants were included in the gram-negative meta-analysis. Notably, 158 included studies with a total of 22,994 participants reported the *total* number of gram-negative bacteria isolated from all participants but did not report the number of participants who had ≥1 gram-negative bacterial species isolated and therefore could not be included in the gram-negative meta-analysis.

### Included studies population characteristics and quality

Included studies and population characteristics are summarized in Table 1, and detailed characteristics are in supplemental tables 6 (*P. aeruginosa* studies) and 7 (gram-negative bacilli of any species studies). The included studies’ quality and population characteristics were similar across outcomes (psaDFI and gnDFI). Below we highlight the studies included in the psaDFI meta-analysis because it contained the largest sample.

**Table 1:** study characteristics by solar climate zones and region.

|  | Overall | Tropical | Subtropical | Temperate | N. America<br>W. Europe | N. Africa<br>Asia |
| --- | --- | --- | --- | --- | --- | --- |
| ***Pseudomonas aeruginosa*** |  |  |  |  |  |  |
| Number of studies | 246 | 80 | 73 | 93 | 46 | 165 |
| Number of participants | 36,788 | 11,106 | 9,876 | 15,806 | 9,068 | 23,687 |
| **Demographics** |  |  |  |  |  |  |
| Complete – studies | 146 (59) | 36 (45) | 41 (56) | 69 (74) | 33 (72) | 91 (55) |
| Complete – participants | 64 (26) | 30 (36) | 22 (30) | 12 (14) | 6 (13) | 52 (32) |
| Proportion male, median (IQR) | 67 (60-75) | 65 (58-73) | 67 (61-76) | 67 (61-75) | 72 (66-79) | 66 (59-74) |
| Mean/median age, median (IQR), years | 60 (57-63) | 58 (56-61) | 57 (53-61) | 61 (59-64) | 62 (56-66) | 60 (57-62) |
| **Recruitment** |  |  |  |  |  |  |
| Convenience or missing - studies | X | 37 (46) | 32 (44) | 25 (27) | 6 (13) | 81 (49) |
| Consecutive or missing- participants | 12842 (35) | 5707 (51) | 4950 (50) | 2185 (14) | 281 (3) | 11822 (50) |
| Outpatient only – studies | 31 (13) | 11 (14) | 10 (14) | 10 (11) | 8 (17) | 16 (10) |
| Outpatient only – participants | 3216 (9) | 1463 (13) | 928 (9) | 735 (5) | 580 (6) | 1796 (8) |
| **Antibiotic exposure** |  |  |  |  |  |  |
| No data – studies | 223 (91) | 77 (96) | 64 (88) | 82 (88) | 39 (85) | 150 (91) |
| No data – participants | 33579 (91) | 10841 (98) | 8734 (88) | 14004 (89) | 8158 (90) | 21427 (90) |
| Antibiotic exposure $\leq 3$ mo. before microbiological test among participants with available data | 1040/3209 (32) | 0/265 (0) | 210/1142 (18) | 830/1802 (46) | 468/910 (51) | 572/2260 (25) |
| **Diabetic foot infection severity** |  |  |  |  |  |  |
| No data – studies | 131 (53) | 40 (50) | 41 (56) | 50 (54) | 28 (61) | 88 (53) |
| No data – participants | 19694 (54) | 6297 (57) | 5230 (53) | 8167 (52) | 5559 (61) | 12515 (53) |
| Moderate-severe infection prevalence among participants with available data | 13143/17094 (77) | 3885/4809 (81) | 3880/4646 (83) | 5378/7639 (70) | 2171/3509 (62) | 8883/11172 (80) |
| **Gram negative of any species** |  |  |  |  |  |  |
| Number of studies | 55 | 14 | 15 | 26 | 19 | 32 |
| Number of participants | 6,079 | 1,124 | 2,072 | 2,883 | 2658 | 2920 |
| **Demographics** |  |  |  |  |  |  |
| Complete – studies | 39 (71) | 5 (36) | 9 (60) | 25 (96) | 16 (84) | 20 (63) |
| Complete – participants | 4450 (67) | 243 (22) | 1642 (80) | 2865 (99) | 2571 (97) | 1817 (62) |
| Proportion male, median (IQR) | 69 (64-76) | 68 (64-72) | 68 (66-77) | 70 (64-78) | 75 (67-79) | 68 (67-74) |
| Mean/median age, median (IQR), years | 60 (56-63) | 55 (50-58) | 57 (53-65) | 61 (58-63) | 63 (56-64) | 59 (53-61) |
| **Recruitment** |  |  |  |  |  |  |
| Convenience or missing - studies | 14 (25) | 5 (36) | 3 (10) | 6 (23) | 2 (11) | 12 (37) |
| Consecutive or missing- participants | 837 (14) | 335 (30) | 214 (10) | 288 (10) | 17 (<1) | 799 (27) |
| Outpatient only – studies | 50 (91) | 14 (100) | 14 (93) | 22 (85) | 16 (84) | 30 (94) |
| Outpatient only – participants | 402 (7) | 128 (11) | 150 (7) | 124 (4) | 66 (2) | 336 (11) |
| **Antibiotic exposure** |  |  |  |  |  |  |
| No data – studies | 50 (91) | 14 (100) | 14 (93) | 22 (85) | 16 (84) | 30 (94) |
| No data – participants | 4857 (80) | 1124 (100) | 1754 (85) | 1979 (69) | 2059 (77) | 2297 (79) |
| Antibiotic exposure $\leq 3$ mo. before microbiological test among participants with available data | 432/1222 (35) | No data | 183/318 (58) | 244/904 (27) | 371/599 (62) | 56/623 (9) |
| **Diabetic foot infection severity** |  |  |  |  |  |  |
| No data – studies | 27 (49) | 6 (43) | 9 (60) | 12 (46) | 10 (53) | 16 (50) |
| No data – participants | 2686 (44) | 418 (37) | 1258 (61) | 1010 (35) | 1289 (48) | 1157 (40) |
| Moderate-severe infection prevalence among participants with available data | 2699/3393 (80) | 644/706 (91) | 767/814 (94) | 1288/1873 (69) | 1008/1369 (74) | 1359/1763 (77) |

All data is presented as number (%) unless otherwise specified. Percentages rounded to the nearest integer and sum of column percentages may not add to 100.
Abbreviations: mo., month.

Among the 246 studies included in the *P. aeruginosa* meta-analysis, 146 (59%) contained complete demographic data. Most participants were male [67% (IQR 60-75%)] and the median mean/median age of the study populations was 60 years old (IQR 57-63). The population demographic characteristics were similar when stratified by climate zone or region. Most studies recruited consecutive participants [148(60%)], but 94 (38%) recruited participants by convenience sampling or did not report their recruitment methods. The proportion of studies that recruited by convenience sampling or had missing recruitment methods was higher in tropical settings [37(46%)] and subtropical settings [32(44%)] compared to temperate settings [25(27%)]. The proportion of studies that recruited by convenience sampling or had missing recruitment methods was lower among studies conducted in North America/Western Europe [6(13%)] compared to North Africa/Asia [81(49%)]. Most studies included no data regarding antibiotic exposure before microbiological sampling [223(91%)] or DFI severity [131 (53%)] with similar missingness rates across climate zones and regions.

### Pooled prevalence of gram-negative and P. aeruginosa diabetic foot infections

Forest plots with the pooled prevalence psaDFIs and gnDFIs stratified by climate zones and regions are shown in supplemental figure 1. We summarize the random effects models point estimates and 95% confidence intervals (95%CI) in Figure 3 given the Forest plots large size. There was substantial between-study heterogeneity, demonstrated by high I^2^ values. The overall psaDFI funnel plot was asymmetric (Egger’s test p<.001) and the overall gnDFI funnel plot was symmetric (Egger’s test p=.07) (supplemental figure 2). Due to the psaDFI plot asymmetry, we built subgroup plots for each climate zone. The tropical and subtropical plots were asymmetric (Egger’s test p<.001) due to an overrepresentation of studies with low psaDFI prevalence, but the temperate plot was symmetric (Egger’s test p=.54).

**Figure 3:**
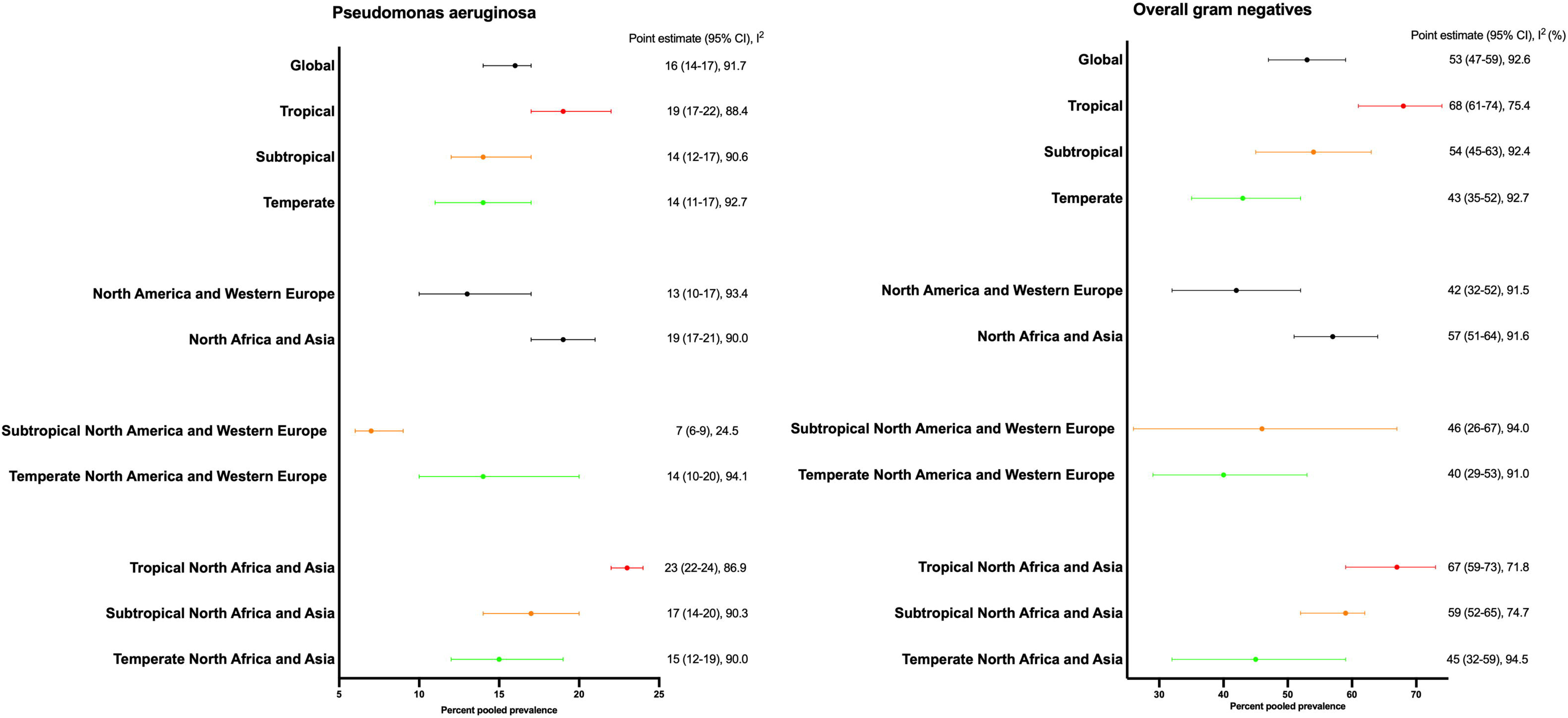
random effects meta-analysis of the pooled *P. aeruginosa* and overall gram negative diabetic foot infection prevalence stratified by solar climate zone and region.

The global pooled prevalence of psaDFIs was 16% (95%CI 14-17%) and gnDFIs was 53% (95%CI 47-59%). The point estimate and 95%CI in tropical settings of psaDFIs [19% (95%CI 17-22%)] and gnDFIs [68% 95%CI 61-74)] was higher compared to temperate settings with little 95%CI overlap for psaDFIs [14% (95%CI 11-17%)] and no 95%CI overlap for gnDFIs [43% (95%CI 35-52%)]. The subtropical psaDFI and gnDFIs pooled prevalences 95%CIs largely overlapped with the 95%CIs of tropical and temperate settings.

The pooled prevalence of psaDFIs and gnDFIs was higher in studies conducted in Asia/North Africa [psaDFI 19% (95%CI 17-21%) and gnDFI 57% (95%CI 51-64%)] compared to studies conducted in North America/Western Europe [psaDFI 13% (95%CI 10-17%) and gnDFI 42% (95%CI 32-52%)]. However, the pooled prevalences of psaDFIs and gnDFIs in temperate zones of Asia/North Africa were similar to those in temperate zones of North America/Western Europe. Within these regions, the pooled prevalence of psaDFIs and gnDFIs followed the same pattern of those observed globally (i.e., higher prevalence in tropical settings) except for psaDFIs in North America/Western Europe, which were more prevalent in temperate settings [14% (95%CI 10-20)] compared to subtropical settings [7% 95%CI (6-9)]. A subgroup analysis comparing studied published between 2010-2015 *vs* 2016-2020 showed similar psaDFI and gnDFI pooled prevalences globally and stratified by climate zones and regions (supplemental table 9).

### Meta-regression

The meta-regression models did not explain between-study heterogeneity, as demonstrated by low R^2^ values (supplemental tables 10-11). The subgroup analysis of studies from North Africa/Asia suggested gnDFI and psaDFI prevalence were associated with climate zones in that region. For example, adjusting for climate and study setting the proportion of gnDFIs and psaDFIs were higher in tropical zones compared to temperate zones [0.89 (95%CI 0.36-1.41)] percentage points for gnDFIs and 0.41 (95%CI 0..03-0.80) percentage points for psaDFIs)].

## DISCUSSION

We performed a comprehensive review to investigate if psaDFIs and gnDFIs are more prevalent in tropical and subtropical zones compared to temperate zones and in North Africa/Asia compared to North America/Western Europe. We found 263 studies published in 13 languages, representing data from 51 countries, and our key findings are that the psaDFIs and gnDFIs are likely more prevalent in tropical settings compared to temperate zones given little or no overlap in the pooled prevalence 95%CI. Conversely, there were no substantial differences in the prevalence of these infections between tropical and subtropical, and subtropical and temperate zones. Additionally, we found that climate zones better discriminate the prevalence of these infections compared to the guideline-proposed regional differences. Importantly, studies generally lacked key data needed to understand if the prevalence differences are related to characteristics other than climate (e.g., antibiotic exposure) and there was substantial between-study heterogeneity within climate zones and regions.

The association between warmer climate and gram-negative infections has been demonstrated in other infections, including bacteremia^15^ and infectious diarrhea^16^, and could be related to climate’s effect on the microbiome^17^ and/or bacterial survival in the environment^18^. In the United States Veterans Affairs healthcare system, Winski *et al* found that psaDFI prevalence varied by International Energy Conservation Code-defined climate zones and ranged from 11.6% in hot humid zones to 6.2% in very cold zones^19^. To our knowledge, this is the first study to investigate psaDFI and gnDFI prevalences globally stratified by climate. Published systematic reviews of DFI microbiology globally stratified their findings by Gross National Income^20^ or geographical regions^21,22^. Macdonald *et al* reviewed studies reporting on DFI microbiology published between the queried database inceptions and 2019^20^. They excluded studies not published in English, did not search WHO databases, and found 112 studies representing 16,159 participants. They did not report patient-level overall gnDFI prevalence but found that the proportion of bacterial isolates that were gram negative was lower in high-income countries (37.6%) compared to upper-middle and low-middle income countries (59.6%) – no studies from low-income countries were included. Qu *et al* reviewed DFI studies published between 2000-2020 and, in addition to PubMed, MEDLINE, and Web of Science, queried several Chinese databases^22^. They found 359 studies representing 56,292 participants - 68% of studies and participants were from China and few studies were from Africa (6), Europe (21), or the Americas (17). They also did not report patient-level overall gnDFI prevalence but found that the proportion of bacterial isolates that were gram negative was 27.3% in the Americas, 35.5% in Europe, 40.4% in Africa, and 45.9% in Asia. Garousi *et al* reviewed studies that reported on psaDFI prevalence published between 2000-2022 and found that, among the 100 included studies, the psaDFI prevalence was 16.6% globally, 18.5% Asia, 16.3% in Africa, and 11.1% in Western countries^21^. Altogether, the literature indicates there are geographical differences in DFI microbiology, and these differences may by partially climate related. Other potential explanations for DFI microbiology variation include differences in footwear use and religion-related foot washing practices.

Our review uncovered several limitations of the DFI literature. First, we could not separate the effect of climate on DFI microbiology from those of other well-established risk factors for psaDFIs and gnDFIs^5^, due to the paucity of data regarding infection severity and prior antibiotic exposure among included studies. Second, most studies reported the total number of gram-negative bacteria as the numerator and total number of bacteria as the denominator. As DFIs are often polymicrobial^6,20^, this impairs our ability to understand patient-level gnDFI prevalence. Third, despite having no language limitation and searching WHO databases, we found studies from only 35 studies outside Asia/North Africa/North America/Western Europe. We need well-designed geographically representative cohort studies that contain patient-level data regarding the contribution of location and clinical factors on DFI microbiological profile and treatment outcomes to produce data that can better guide antimicrobial therapy. We also identified potential for publication bias regarding psaDFI prevalence and many studies recruited a convenience participant sample or did not report their sampling methods.

As a strength, the demographics of included studies was representative of the diabetic foot disease epidemiology globally, which affects mostly men and persons aged 50-69^1^. Our study also has limitations. First, we excluded multisite studies when they included participants from >1 climate zone and did not stratify results by climate zone. Second, we included studies over a 10-year period. However, the psaDFI and gnDFI prevalences were similar for studies published between 2010-2015 *vs* 2016-2020. Third, we did not separate bone from soft tissue sample results. Fourth, while we used widely accepted definitions of tropical and subtropical settings, there are no universally accepted definitions of these climate zones^9^. Lastly, the includes studies are all subnational and come from healthcare facility-level reports and therefore may miss community-level DFIs.

In conclusion, we report the first meta-analysis investigating the associations between climate and DFI microbiology and found a higher psaDFI and gnDFI prevalence in tropical zones compared to temperate zones. We uncovered limitations of the DFI literature and suggests ways to improve our understanding of this topic. Better understanding of the risk factors for psaDFI and gnDFI can improve antimicrobial utilization.

## Supporting information

Search Strategy

COI forms

Prospero registration

PRISMA statement

Raw data

## Data Availability

All data produced are available online in the supplemental files.

## DATA SHARING

All raw data and analysis R code are available in the supplemental files

---

**Supplemental figure 1A:**
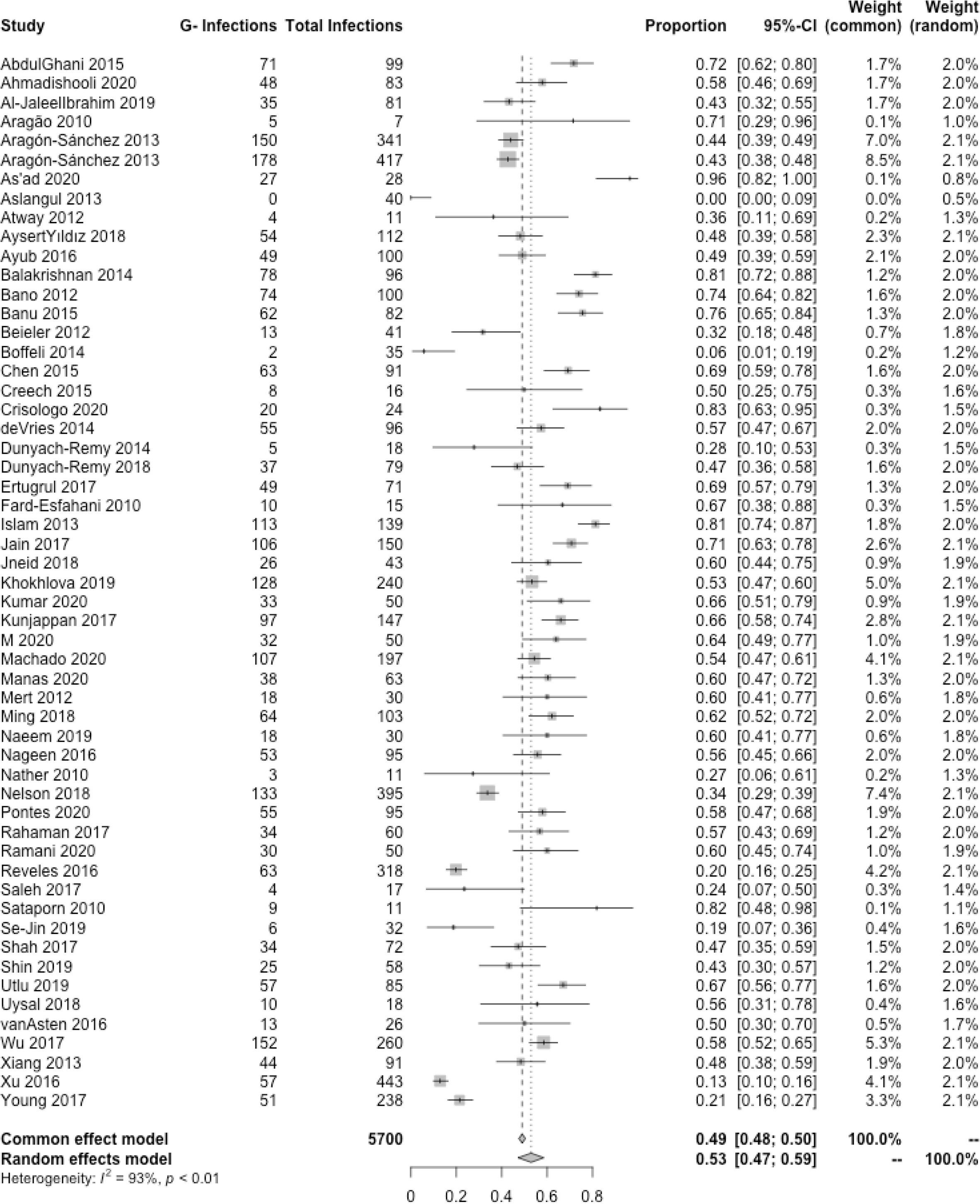
Forest plot of studies reporting on overall gram negative diabetic foot infection prevalence.

**Supplemental figure 1B:**
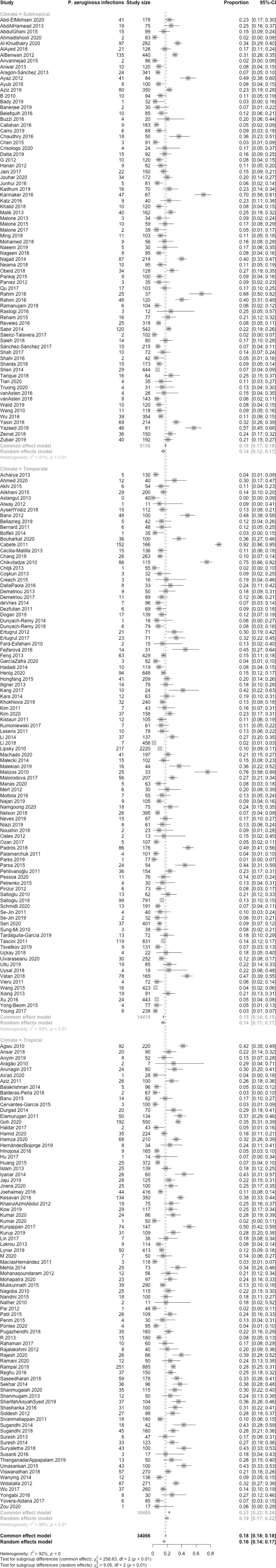
Forest plot of studies reporting on P. aeruginosa foot infection prevalence.

**Supplemental figure 2:**
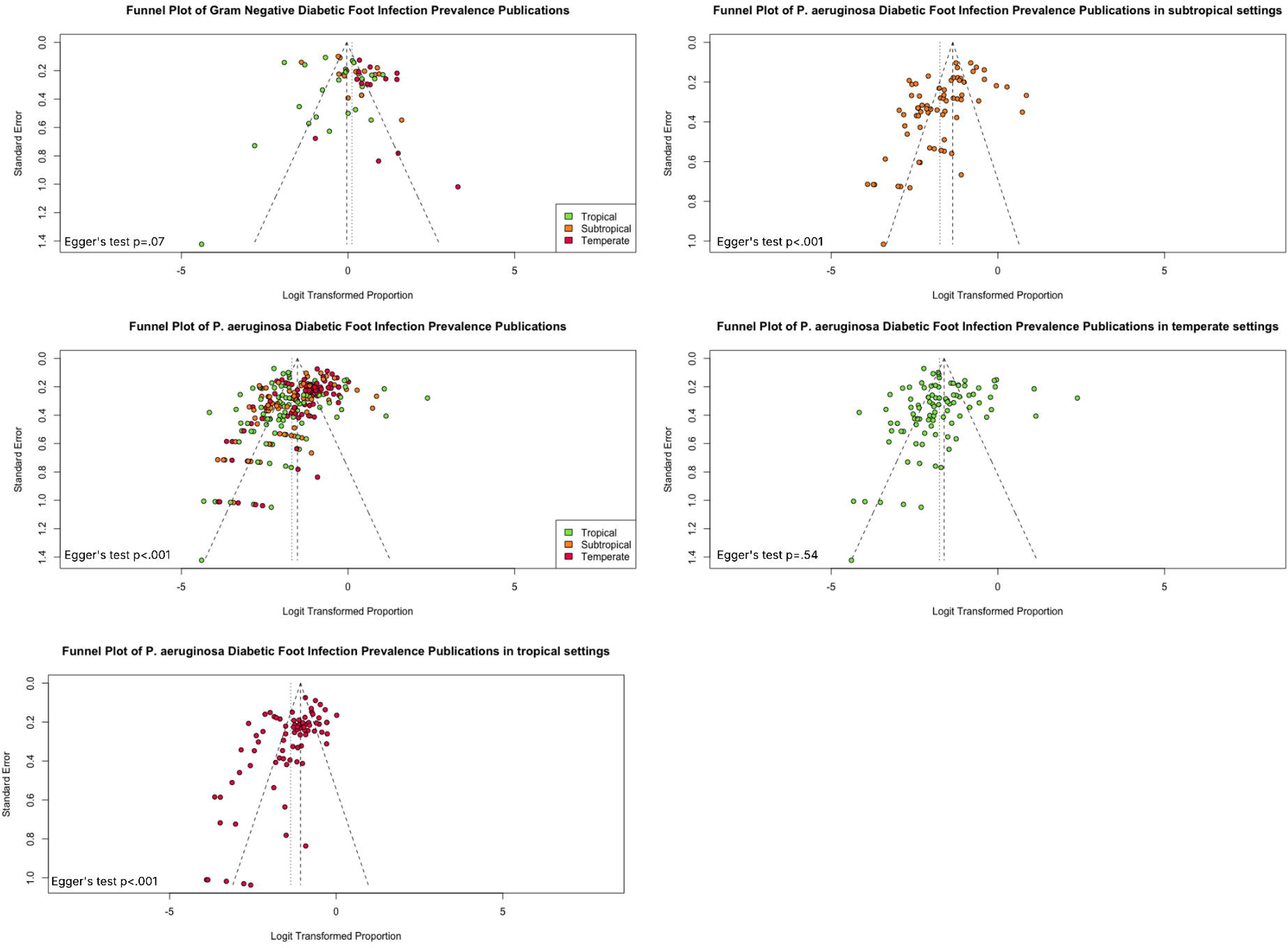
Funnel plots.

