## Supplementary material for "Global gram-negative diabetic foot infection prevalence and its associations with climate: a systematic review and meta-analysis": Search Strategy

**All searches were conducted December 15, 2020**

**PubMed:**

1. ("Diabetic Foot"[MeSH Terms] AND ("infections"[MAJR] OR "soft tissue infect*"[Text Word] OR "Osteomyelitis"[Text Word] OR ulcer*[Text Word])) OR "diabetic foot infect*"[Text Word]

2. "Foot"[MeSH Terms] AND ("Diabetes Complications"[MAJR] OR “diabetes mellitus/complications”[MAJR])

3. (("diabetic foot"[Text Word] OR (“diabetes”[TW] AND “foot”[TW])) AND (infect[Text Word] OR infected[TW] OR infection[TW] OR infections[TW] OR infecting[TW] OR ulcer*[TW] OR osteomyelitis[TW] OR “soft tissue infect*”[TW])) OR DFI[TW] OR DFU[TW]

4. #1 OR #2 OR #3

5. "Bacterial Typing Techniques"[MeSH Terms] OR "bacterial typing"[Text Word] OR "bacterial identif*"[Text Word] OR "microbiology"[Text Word] OR "microbiology"[MeSH Subheading] OR "diagnosis"[Text Word] OR "diagnosis" [Subheading:NoExp] OR "isolation and purification"[MeSH Subheading] OR pathogen*[Text Word] OR "culture"[Text Word] OR (bacteria*[TW] AND classification[TW])

6. ((#4 AND #5) AND (“Journal article”[PTYP] OR “journal article”[TW]) AND "2010/01/01"[PDat] : "3000/12/31"[PDat]) NOT (“pubmed books”[Filter] OR “editorial”[PTYP] OR “personal narrative”[PTYP])

**Embase**

1. ‘Diabetic Foot’/exp AND ('infection'/exp/mj OR ‘soft tissue infection*’:ti,ab,kw OR ulcer*:ti,ab,kw)

2. 'diabetic foot osteomyelitis'/exp

3. (‘diabetic foot*’:ti,ab,kw)

4. (‘diabetic foot*’ NEAR/3 (infect* OR ulcer* OR osteomyelitis)) OR ((diabetes NEAR/3 foot) AND (infect* OR ulcer* OR osteomyelitis):ti,ab,kw) OR DFI:ti,ab,kw OR DFU:ti,ab,kw

5. #1 OR #2 OR #3 OR #4

### 6. 'bacterium identification'/exp OR bacteria*:ti,ab,kw OR (bacteria* NEAR/3 (identif* OR infect* OR classification OR diagnose OR diagnosis)) OR microbiology:ti,ab,kw OR 'microbiology'/exp/mj OR 'diagnosis'/exp/mj OR diagnosis:ti,ab,kw OR 'isolation and purification'/exp OR pathogen*:ti,ab,kw OR (identif* NEAR/3 pathogen*) OR Culture:ti,ab,kw

### 7. #5 AND #6

### #8 #7 AND (2010:py OR 2011:py OR 2012:py OR 2013:py OR 2014:py OR 2015:py OR 2016:py OR 2017:py OR 2018:py OR 2019:py OR 2020:py) AND ([article]/lim OR [article in press]/lim OR [letter]/lim OR [review]/lim) NOT (([animals]/lim OR ‘animals’/exp OR animal*:ti,ab,kw) NOT ([humans]/lim OR 'human'/exp OR human*:ti,ab,kw))

### Web of Science:

### 1. (TS=("diabetic foot" NEAR/3 (infect* OR ulcer* OR osteomyelitis)) OR TS=((diabetes AND foot* NEAR/3 (infect* OR ulcer* OR osteomyelitis)))

### 2. TS= (bacteri* OR microbio* OR culture* OR pathogen* NEAR/3 (diagnosis OR diagnose OR identi* OR classif OR etiology) )

### 3. #1 AND #2

### 4. (#3)  AND DOCUMENT  TYPES: (Article OR Early Access OR Letter OR Review)  Indexes=SCI-EXPANDED, SSCI, A&HCI, CPCI-S, CPCI-SSH, BKCI-S, BKCI-SSH, ESCI, CCR-EXPANDED, IC Timespan=2010-2020

### WHO databases, specifically: AIM, IMSEAR, IMEMR, LILACS WPRIM

### (tw: (diabetes foot infection)) OR (tw:(diabetic foot* bacteri*)) OR (tw:(diabetic foot* microbiolog*)) OR (tw:(diabetic foot* pathogen*))
