## Supplemental tables for "Global gram-negative diabetic foot infection prevalence and its associations with climate: a systematic review and meta-analysis"

Supplemental table **1**: Study latitude and longitude classification system

| **Level 1:** One hospital or clinic listed | Use the hospital or clinic address latitude and longitude |
| --- | --- |
| **Level 2:** Multiple hospitals or clinics within one city | Use the first hospital or clinic listed in the authors affiliation section address latitude and longitude |
| **Level 3:** Multiple hospitals or clinics within different cities | The location where participants are receiving care, if indicated in the study  OR  Use the most populous city latitude and longitude |
| **Level 4:** A country, region, state, or city is given or there is no location given AND all authors are from the same city | Use authors primary affiliation latitude and longitude |
| **Level 5:** A country, region, or state is given AND researchers are from different cities | Use the latitude and longitude of the first author’s primary affiliation |

Supplemental table 2: study definitions North Africa/Asia and North America/Western Europe.

Based on the WHO “Standard country or area codes for statistical use (M49)”

### Northern America

### Canada

### Greenland

### Bermuda

### Saint Pierre and Miquelon

### United States

### Western Europe – the WHO Western and Southern European regions were classified together as Western Europe.

### Albania

### Andorra

### Bosnia and Herzegovina

### Croatia

### Gibraltar

### Greece

### Holy See

### Italy

### Malta

### Montenegro

### North Macedonia

### Portugal

### San Marino

### Serbia

### Slovenia

### Spain

### Austria

### Belgium

### France

### Germany

### Liechtenstein

### Luxembourg

### Monaco

### Netherlands

### Switzerland

### Northern Africa

### Algeria

### Egypt

### Libya

### Morocco

### Sudan

### Tunisia

### Western Sahara

### Asia

### Kazakhstan

### Kyrgyzstan

### Tajikistan

### Turkmenistan

### Uzbekistan

### China

### Hong Kong

### Democratic People's Republic of Korea

### Japan

### Mongolia

### Republic of Korea

### Brunei Darussalam

### Cambodia

### Indonesia

### Lao People's Democratic Republic

### Malaysia

### Myanmar

### Philippines

### Singapore

### Thailand

### Timor-Leste

### Viet Nam

### Afghanistan

### Bangladesh

### Bhutan

### India

### Iran (Islamic Republic of)

### Maldives

### Nepal

### Pakistan

### Sri Lanka

### Armenia

### Azerbaijan

### Bahrain

### Cyprus

### Georgia

### Iraq

### Israel

### Jordan

### Kuwait

### Lebanon

### Oman

### Qatar

### Saudi Arabia

### State of Palestine

### Syrian Arab Republic

### Türkiye

### United Arab Emirates

### Yemen

**Supplemental table 3: study risk of bias and data quality assessment**

| Demographic data | |
| --- | --- |
| Complete data | A-The proportion of males and females *AND*  B- The study mean or median population age were available |
| Incomplete data | A-The proportion of males and females *OR*  B- The study mean or median population age were available |
| No data | None of the above reported |
| Participant recruitment | |
| Consecutive | Consecutive participants that met the original study criteria enrolled |
| Random | Random subset of the persons with diabetic foot infection enrolled |
| Convenience or missing | Convenience sample or recruitment methods unclear |
| Antibiotic exposure | |
| Complete data | A- The proportion of the study population exposed to antibiotics within a defined period prior to microbiological testing was reported *AND*  B- The microbiological results were stratified by prior antibiotic exposure |
| Incomplete data | A-The proportion of the study population exposed to antibiotics within a defined period prior to microbiological testing was reported *BUT*  B-The microbiological results were NOT stratified by prior antibiotic exposure |
| No data | The proportion of the study population exposed to antibiotics within a defined period prior to microbiological testing was NOT reported |
| Diabetic foot infection severity | |
| Complete data | A- The proportion of patients with severe, moderate, or mild infection per the PEDIS/IDSA, Wagner, and/or University of Texas classification systems was reported *AND*  B- The microbiological results were stratified by infection severity |
| Incomplete data | A- The proportion of patients with severe, moderate, or mild infection per the PEDIS/IDSA, Wagner, and/or University of Texas classification systems was reported *BUT*  B- The microbiological results were NOT stratified by infection severity |
| No data | The proportion of patients with severe, moderate, or mild infection per the PEDIS/IDSA, Wagner, and/or University of Texas classification systems was NOT reported |

### Supplemental table 4: Diabetic foot infection severity classification systems

| Classification system | Severity | Re-classification for the meta-analysis |
| --- | --- | --- |
| IDSA/IWGDF | | |
| 1 | Mild | Mild |
| 2 | Moderate | Moderate-Severe |
| 3- | Severe | Moderate-Severe |
| Meggitt-Wagner | | |
| 0 | Healed or pre-ulcerative wound | Mild |
| 1 | Superficial ulcer (involves epidermis, dermis, or subcutaneous tissue) | Mild |
| 2 | Deep ulcer (*reaches tendon, bone, or joint capsule)* | Mild |
| 3 | Deeper tissues are involved and there is abscess formation, osteomyelitis, or tendinitis | Moderate-Severe |
| 4 | Limited gangrene (*part of the foot)* | Moderate-Severe |
| 5 | Extensive gangrene (*whole foot*) | Moderate-Severe |
| University of Texas ^1^ | | |
| 0 | Pre-ulcerative or post-ulcerative healed wound | Mild |
| 1 | Superficial ulcer not involving tendon, capsule, or bone | Mild |
| 2 | Wound penetrating to tendon or capsule | Moderate-Severe |
| 3 | Wound penetrating to bone or joint | Moderate-Severe |

### 1-The University of Texas wound grades are further subclassified in 4 stages: (a) No infection or ischemia, (b) Infection present (c), Ischemia present, and (d) infection and ischemia present

Supplemental table 5: included studies publication language and country

| **Country** | **Studies** | **Total population** | **Region assigned** |
| --- | --- | --- | --- |
| Algeria | 1 | 117 | Asia/North Africa |
| Argentina | 1 | 72 | Other |
| Australia | 3 | 511 | Other |
| Bangladesh | 1 | 67 | Asia/North Africa |
| Brazil | 4 | 204 | Other |
| Brunei | 1 | 75 | Asia/North Africa |
| Cameron | 1 | 30 | Other |
| China | 22 | 4822 | Asia/North Africa |
| Cote d’Ivoire | 1 | 241 | Other |
| Czech Republic | 1 | 31 | Other |
| Egypt | 6 | 643 | Asia/North Africa |
| France | 6 | 331 | Western Europe/North America |
| Georgia | 2 | 220 | Other |
| Germany | 1 | 78 | Western Europe/North America |
| Greece | 3 | 196 | Western Europe/North America |
| Guyana | 1 | 109 | Other |
| India | 59 | 6513 | Asia/North Africa |
| Indonesia | 2 | 45 | Asia/North Africa |
| Iran | 13 | 1049 | Asia/North Africa |
| Iraq | 4 | 762 | Asia/North Africa |
| Israel | 1 | 61 | Asia/North Africa |
| Italy | 2 | 1328 | Western Europe/North America |
| Kuwait | 1 | 440 | Asia/North Africa |
| Lebanon | 2 | 484 | Asia/North Africa |
| Malaysia | 6 | 2224 | Asia/North Africa |
| Mexico | 6 | 767 | Other |
| Morocco | 1 | 157 | Asia/North Africa |
| Netherlands | 1 | 110 | Western Europe/North America |
| Nicaragua | 1 | 34 | Other |
| Nigeria | 2 | 272 | Other |
| Pakistan | 17 | 2250 | Asia/North Africa |
| Peru | 1 | 88 | Other |
| Poland | 2 | 163 | Other |
| Portugal | 5 | 678 | Western Europe/North America |
| Romania | 3 | 397 | Other |
| Russia | 3 | 624 | Other |
| Saudi Arabia | 4 | 315 | Asia/North Africa |
| Singapore | 2 | 111 | Asia/North Africa |
| South Korea | 8 | 650 | Asia/North Africa |
| Spain | 7 | 1367 | Western Europe/North America |
| Sudan | 3 | 760 | Asia/North Africa |
| Switzerland | 1 | 22 | Western Europe/North America |
| Syria | 1 | 100 | Asia/North Africa |
| Taiwan | 1 | 98 | Asia/North Africa |
| Thailand | 1 | 11 | Asia/North Africa |
| Trinidad and Tobago | 1 | 139 | Other |
| Tunisia | 1 | 112 | Asia/North Africa |
| Turkey | 15 | 2401 | Asia/North Africa |
| Ukraine | 2 | 131 | Other |
| United Kingdom | 6 | 789 | Western Europe/North America |
| United States | 19 | 4811 | Western Europe/North America |
| **Publication language** | **Studies** |  |  |
| Arabic | 1 |  |  |
| Chinese | 11 |  |  |
| Czech | 1 |  |  |
| English | 223 |  |  |
| Farsi | 4 |  |  |
| French | 1 |  |  |
| Korean | 4 |  |  |
| Portuguese | 2 |  |  |
| Romanian | 1 |  |  |
| Russian | 4 |  |  |
| Spanish | 5 |  |  |
| Ukrainian | 2 |  |  |
| Turkish | 4 |  |  |

Supplemental table 6: extended study characteristics by solar climate zones and regions – *Pseudomonas aeruginosa* studies

|  | Overall | Tropical | Subtropical | Temperate | N. America and W. Europe | N. Africa and Asia |
| --- | --- | --- | --- | --- | --- | --- |
|  | 246 studies  36,788 participants | 80 studies  11,106 participants | 73 studies  9,876 participants | 93 studies  15,806  participants | 46 studies  9,068  participants | 165 studies  23,687  participants |
| Demographics |  |  |  |  |  |  |
| Complete data – studies | 146 (59) | 36 (45) | 41 (56) | 69 (74) | 33 (72) | 91 (55) |
| Incomplete data – studies | 64 (26) | 30 (36) | 22 (30) | 12 (14) | 6 (13) | 52 (32) |
| No data – studies | 37 (15) | 14 (18) | 10 (14) | 12 (14) | 7 (15) | 22 (13) |
| Complete data – participants | 22165 (60) | 3954 (37) | 5979 (61) | 12232 (77) | 6747 (74) | 12700 (54) |
| Incomplete data – participants | 9263 (25) | 5575 (50) | 2441 (25) | 1247 (8) | 643 (7) | 8083 (34) |
| No data – participants | 5360 (15) | 1577 (14) | 1456 (15) | 2327 (15) | 1678 (19) | 2904 (12) |
| Recruitment |  |  |  |  |  |  |
| Consecutive - studies | 148 (60) | 39 (49) | 41 (56) | 68 (73) | 40 (87) | 80 (48) |
| Random - studies | 4 (2) | 4 (5) | 0 | 0 | 0 | 4 (2) |
| Convenience or missing - studies | 94 (38) | 37 (46) | 32 (44) | 25 (27) | 6 (13) | 81 (49) |
| Consecutive - participants | 23499 (64) | 4916 (44) | 4926 (50) | 13621 (86) | 8787 (97) | 11382 (48) |
| Random - participants | 483 (1) | 483 (4) | 0 | 0 | 0 | 0 |
| Convenience or missing - participants | 12842 (35) | 5707 (51) | 4950 (50) | 2185 (14) | 281 (3) | 11822 (50) |
| Study setting |  |  |  |  |  |  |
| Outpatient only - studies | 31 (13) | 11 (14) | 10 (14) | 10 (11) | 8 (17) | 16 (10) |
| Inpatient/emergency - studies | 169 (69) | 50 (63) | 55 (75) | 64 (69) | 31 (67) | 113 (68) |
| Missing - studies | 46 (19) | 19 (24) | 8 (11) | 19 (20) | 7 (15) | 36 (22) |
| Outpatient only - participants | 3216 (9) | 1463 (13) | 928 (9) | 735 (5) | 580 (6) | 1796 (8) |
| Inpatient/emergency - participants | 26516 (72) | 7217 (65) | 7184 (73) | 12115 (77) | 7571 (83) | 15962 (67) |
| Missing - participants | 7146 (19) | 2426 (22) | 1764 (18) | 2956 (19) | 917 (10) | 5929 (25) |
| Antibiotic exposure |  |  |  |  |  |  |
| Complete data – studies | 13 (5) | 2 (3) | 7 (10) | 4 (4) | 3 (7) | 9 (6) |
| Incomplete data – studies | 10 (4) | 1 (1) | 2 (3) | 7 (8) | 4 (9) | 6 (4) |
| No data – studies | 223 (91) | 77 (96) | 64 (88) | 82 (88) | 39 (85) | 150 (91) |
| Complete data – participants | 1652 (4) | 150 (1) | 763 (7) | 739 (5) | 206 (2) | 1407 (6) |
| Incomplete data – participants | 1557 (4) | 115 (1) | 379 (4) | 1063 (7) | 704 (8) | 853 (4) |
| No data – participants | 33579 (91) | 10841 (98) | 8734 (88) | 14004 (89) | 8158 (90) | 21427 (90) |
| Antibiotic exposure 3 months before microbiological test – among participants with available data | 1040/3209  (32) | 0/265  (0) | 210/1142  (18) | 830/1802  (46) | 468/910  (51) | 572/2260  (25) |
| Diabetic foot infection severity |  |  |  |  |  |  |
| Complete data – studies | 12 (5) | 5 (6) | 4 (5) | 3 (3) | 2 (4) | 8 (5) |
| Incomplete data – studies | 103 (42) | 35 (44) | 28 (38) | 40 (43) | 16 (35) | 69 (42) |
| No data – studies | 131 (53) | 40 (50) | 41 (56) | 50 (54) | 28 (61) | 88 (53) |
| Complete data – participants | 1927 (5) | 686 (6) | 906 (9) | 335 (2) | 128 (1) | 1351 (6) |
| Incomplete data – participants | 15167 (41) | 4123 (37) | 3740 (38) | 7304 (46) | 3381 (37) | 9821 (42) |
| No data – participants | 19694 (54) | 6297 (57) | 5230 (53) | 8167 (52) | 5559 (61) | 12515 (53) |
| Moderate-severe infection - among participants with available data | 13143/17094  (77) | 3885/4809  (81) | 3880/4646  (83) | 5378/7639  (70) | 2171/3509  (62) | 8883/11172  (80) |

All data is presented as number (%) unless otherwise specified. Percentages rounded to the nearest integer and sum of column percentages may not add to 100.

Abbreviations: mo., month.

Supplemental table 7: extended study characteristics by solar climate zones and regions – gram-negative infections of any species studies

|  | Overall | Tropical | Subtropical | Temperate | N. America and W. Europe | N. Africa and Asia |
| --- | --- | --- | --- | --- | --- | --- |
|  | 55 studies  6.079 participants | 14 studies  1,124 participants | 15 studies  2,072 participants | 26 studies  2,883 participants | 19 studies  2658  participants | 32 studies  2920 participants |
| Demographics |  |  |  |  |  |  |
| Complete data – studies | 39 (71) | 5 (36) | 9 (60) | 25 (96) | 16 (84) | 20 (63) |
| Incomplete data – studies | 9 (16) | 5 (36) | 4 (27) | 0 | 1 (5) | 7 (22) |
| No data – studies | 7 (13) | 4 (29) | 2 (13) | 1 (4) | 2 (11) | 5 (16) |
| Complete data – participants | 4450 (67) | 243 (22) | 1642 (80) | 2865 (99) | 2571 (97) | 1817 (62) |
| Incomplete data – participants | 750 (12) | 435 (39) | 315 (15) | 0 | 35 (1) | 576 (19) |
| No data – participants | 579 (10) | 446 (40) | 115 (6) | 18 (1) | 52 (2) | 527 (18) |
| Recruitment |  |  |  |  |  |  |
| Consecutive - studies | 41 (75) | 9 (64) | 12 (80) | 20 (77) | 17 (89) | 20 (63) |
| Random - studies | 0 | 0 | 0 | 0 | 0 | 0 |
| Convenience or missing - studies | 14 (25) | 5 (36) | 3 (10) | 6 (23) | 2 (11) | 12 (37) |
| Consecutive - participants | 5242 (86) | 789 (70) | 1858 (90) | 2595 (90) | 2629 (99) | 2121 (73) |
| Random - participants | 0 | 0 | 0 | 0 | 0 | 0 |
| Convenience or missing - participants | 837 (14) | 335 (30) | 214 (10) | 288 (10) | 17 (<1) | 799 (27) |
| Study setting |  |  |  |  |  |  |
| Outpatient only - studies | 6 (11) | 2 (14) | 1 (6) | 3 (12) | 2 (11) | 4 (12) |
| Inpatient/emergency - studies | 40 (73) | 12 (86) | 12 (80) | 16 (62) | 14 (74) | 22 (69) |
| Missing - studies | 9 (16) | 0 | 2 (13) | 7 (27) | 3 (16) | 6 (19) |
| Outpatient only - participants | 402 (7) | 128 (11) | 150 (7) | 124 (4) | 66 | 336 (11) |
| Inpatient/emergency - participants | 4375 (72) | 996 (89) | 1758 (85) | 1621 (56) | 2122 | 1752 (60) |
| Missing - participants | 1302 (19) | 0 | 164 (8) | 1745 (61) | 470 | 832 (28) |
| Antibiotic exposure |  |  |  |  |  |  |
| Complete data – studies | 2 (4) | 0 | 0 | 2 (7) | 1 (5) | 1 (3) |
| Incomplete data – studies | 3 (5) | 0 | 1 (7) | 2 (7) | 2 (11) | 1 (3) |
| No data – studies | 50 (91) | 14 (100) | 14 (93) | 22 (85) | 16 (84) | 30 (94) |
| Complete data – participants | 576 (9) | 0 | 0 | 576 (20) | 43 (<1) | 533 (18) |
| Incomplete data – participants | 646 (11) | 0 | 318 (15) | 328 (11) | 556 (21) | 90 (3) |
| No data – participants | 4857 (80) | 1124 (100) | 1754 (85) | 1979 (69) | 2059 (77) | 2297 (79) |
| Antibiotic exposure 3 months before microbiological test – among participants with available data | 432/1222  (35) | No data | 183/318 (58) | 244/904 (27) | 371/599  (62) | 56/623  (9) |
| Diabetic foot infection severity |  |  |  |  |  |  |
| Complete data – studies | 5 (9) | 2 (14) | 1 (7) | 2 (8) | 2 (11) | 3 (9) |
| Incomplete data – studies | 23 (42) | 6 (43) | 5 (33) | 12 (46) | 7 (37) | 13 (41) |
| No data – studies | 27 (49) | 6 (43) | 9 (60) | 12 (46) | 10 (53) | 16 (50) |
| Complete data – participants | 502 (8) | 271 (24) | 103 (5) | 128 (4) | 128 (5) | 374 (13) |
| Incomplete data – participants | 2891 (48) | 435 (39) | 711 (34) | 1745 (61) | 1241 (47) | 1389 (48) |
| No data – participants | 2686 (44) | 418 (37) | 1258 (61) | 1010 (35) | 1289 (48) | 1157 (40) |
| Moderate-severe infection - among participants with available data | 2699/3393 (80) | 644/706 (91) | 767/814 (94) | 1288/1873 (69) | 1008/1369  (74) | 1359/1763  (77) |

All data is presented as number (%) unless otherwise specified. Percentages rounded to the nearest integer and sum of column percentages may not add to 100.

Abbreviations: mo., month.

Supplemental table 8: meta-analysis results stratified by studies published between 2010-2015 *vs* 2016-2020

|  | *P. aeruginosa* DFI pooled prevalence % (95%CI)  Studies published between 2010-2015  (103 studies) | *P. aeruginosa* DFI pooled prevalence % (95%CI)  Studies published between 2016-2020  (143 studies) | Gram negative DFI pooled prevalence % (95%CI)  Studies published between 2010-2015  (21 studies) | Gram negative DFI pooled prevalence % (95%CI)  Studies published between 2016-2020  (34 studies) |
| --- | --- | --- | --- | --- |
| Global | 16 (13-19) | 16 (14-18) | 54 (43-65) | 51 (45-58) |
| Solar climate zone |  |  |  |  |
| Tropical | 19 (15-24) | 21 (18-25) | 74 (60-84) | 61 (57-64) |
| Subtropical | 14 (9-19) | 15 (12-19) | 56 (40-71) | 53 (42-63) |
| Temperate | 15 (11-20) | 14 (11-18) | 41 (26-58) | 43 (33-54) |
| Region |  |  |  |  |
| N. Africa/Asia | 17 (14-20) | 19 (16-22) | 66 (56-75) | 53 (46-61) |
| N. America/Western Europe | 15 (8-26) | 11 (8-15) | 36 (22-53) | 45 (32-60) |

Supplemental table 9: gram-negative of any species meta-regression

|  |  |  |  |  | Association between study characteristics and gram-negative diabetic foot infection prevalence (95% CI) | | | | |
| --- | --- | --- | --- | --- | --- | --- | --- | --- | --- |
| Model | I^2^ | R^2^ | Studies included (n) | Studies excluded due to missing data (n) | Tropical | Subtropical | North America/Western Europe | Outpatient recruitment only | Country-level antibiotic consumption  (1000 daily defined doses) |
| **Global** | | | | | | | | | |
| Solar climate | 91% | 20% | 55 | 0 | 1.01  (0.49 – 1.53) | 0.43  (-0.04 – 0.92) | NI | NI | NI |
| Solar climate + recruitment setting | 90% | 18% | 46 | 9 | **0.94**  **(0.42 – 1.46)** | 0.38  (-0.11 – 0.89) | NI | 0  (-0.69-0.67) | NI |
| Region | 92% | 10% | 51 | 0 | NI | NI | **-0.60**  **(-1.07 – 0.13)** | NI | NI |
| Region + recruitment setting | 90% | 12% | 42 | 9 | NI | NI | **-0.58**  **(-1.06 - -0.10)** | -0.06  (-0.77 – 0.65) | NI |
| **North America or Western Europe** | | | | | | | | | |
| Solar climate | 95% | 0% | 19 | 0 | NA | 0.02  (-0.74 – 1.21) | NA | NI | NI |
| Solar climate + recruitment setting | 93% | 15% | 16 | 3 | NA | -0.20  (-1.10 – 0.70) | NA | **-1.78**  **(-3.2 – 0.30)** | NI |
| **Asia or North Africa** | | | | | | | | | |
| Solar climate | 87% | 21% | 32 | 0 | **0.91**  **(0.30-1.51)** | 0.55  (-0.03 – 1.14) | NA | NI | NI |
| Solar climate + recruitment setting | 75% | 34% | 26 | 6 | **0.89**  **(0.36-1.41)** | **0.64**  **(0.11-1.17)** | NA | 0.47  (-0.10 – 1.06) | NI |

Model including median study year recruitment only (I^2^ 93%, R^2^ 1%, 8 studies excluded due to missing data, change in gram-negative DFI prevalence per year of recruitment 0.04 (95%CI = -0.02 – 0.11)

Abbreviations: NA, not applicable; NI, not included

Supplemental table 10: *P. aeruginosa* meta-regression

|  |  |  |  |  | Association between study characteristics and *P. aeruginosa* diabetic foot infection prevalence (95% CI) | | | | |
| --- | --- | --- | --- | --- | --- | --- | --- | --- | --- |
| Model | I^2^ | R^2^ | Studies included (n) | Studies excluded due to missing data (n) | Tropical | Subtropical | North America/Western Europe | Outpatient recruitment only | Country-level antibiotic consumption  (1000 daily defined doses) |
| **Global** | | | | | | | | | |
| Solar climate | 93% | 3% | 246 | 0 | **0.34**  **(0.05 – 0.62)** | -0.01  (-0.29 – 0.28) | NI | NI | NI |
| Solar climate + recruitment setting | 92% | 3% | 200 | 46 | 0.29  (-0.01 – 0.61) | -0.02  (-0.33 – 0.27) | NI | 0.19  (-0.15 – 0.55) | NI |
| Region | 93% | 3% | 211 | 0 | NI | NI | **-0.38**  **(-0.69 - -0.07)** | NI | NI |
| Region + recruitment setting | 92% | 4% | 168 | 43 | NI | NI | **-0.41**  **(-0.73 - -0.09)** | 0.09  (-0.29 – 0.48) | NI |
| **North America or Western Europe** | | | | | | | | | |
| Solar climate | 95% | 4% | 46 | 0 | NA | -0.59  (-1.46 – 0.27) | NA | NI | NI |
| Solar climate + recruitment setting | 92% | 4% | 39 | 7 | NA | -0.58  (-1.34 – 0.17) | NA | -0.17  (-0.97 – 0.61) | NI |
| **Asia or North Africa** | | | | | | | | | |
| Solar climate | 91% | 6% | 165 | 0 | **0.46**  **(0.14 – 0.78)** | 0.12  (-0.20 – 0.45) | NA | NI | NI |
| Solar climate + recruitment setting | 91% | 4% | 129 | 36 | **0.41**  **(0.03 – 0.80)** | 0.08  (-0.29 – 0.46) | NA | 0.09  (-0.35 – 0.53) | NI |

Model including median study year recruitment only (I^2^ 94%, R^2^ 16%, 42 studies excluded due to missing data, change in *P. aeruginosa* DFI prevalence per year of recruitment 0.00 (95%CI = -0.03 – 0.0.4)

Abbreviations: NA, not applicable; NI, not included
