## Supplementary material for "Global gram-negative diabetic foot infection prevalence and its associations with climate: a systematic review and meta-analysis": COI forms

**ICMJE DISCLOSURE FORM**

**Date:** Feb 14, 2025

**Your Name:** Marcos C. Schechter

**Manuscript number (if known):__________________________________________________________________**

**In the interest of transparency, we ask you to disclose all relationships/activities/interests listed below that are**

**related to the content of your manuscript. “Related” means any relation with for-profit or not-for-profit third**

**parties whose interests may be affected by the content of the manuscript. Disclosure represents a commitment**

**to transparency and does not necessarily indicate a bias. If you are in doubt about whether to list a relationship/activity/interest, it is preferable that you do so.**

**The following questions apply to the author’s relationships/activities/interests as they relate to the current**

**manuscript only.**

**The author’s relationships/activities/interests should be defined broadly. For example, if your manuscript pertains**

**to the epidemiology of hypertension, you should declare all relationships with manufacturers of antihypertensive medication, even if that medication is not mentioned in the manuscript.**

**In item #1 below, report all support for the work reported in this manuscript without time limit. For all other items,**

**the time frame for disclosure is the past 36 months.**

|  |  | **Name all entities with whom you have this relationship or indicate none (add rows as needed)** | | **Specifications/Comments**  **(e.g., if payments were made to you or to your institution)** |
| --- | --- | --- | --- | --- |
| **Time frame: Since the initial planning of the work** | | | | |
| 1 | All support for the present manuscript (e.g., funding, provision of study materials, medical writing, article processing charges, etc.)  **No time limit for this item.** | None | |  |
| **Time frame: past 36 months** | | | | |
| 2 | Grants or contracts from any entity (if not indicated in item #1 above). | None |  | |
| 3 | Royalties or licenses | None |  | |
| 4 | Consulting fees | None |  | |
| 5 | Payment or honoraria for lectures, presentations, speakers bureaus, manuscript writing or educational events | None |  | |
| 6 | Payment for expert testimony | None |  | |
| 7 | Support for attending meetings and/or travel | None |  | |
| 8 | Patents planned, issued or pending | None |  | |
| 9 | Participation on a Data  Safety Monitoring Board or Advisory Board | None |  | |
| 10 | Leadership or fiduciary role in other board, society, committee or advocacy group, paid or unpaid | None |  | |
| 11 | Stock or stock options | None |  | |
| 12 | Receipt of equipment, materials, drugs, medical writing, gifts or other services | None |  | |
| 13 | Other financial or non-financial interests | None |  | |

**Please place an “X” next to the following statement to indicate your agreement:**

**X	I certify that I have answered every question and have not altered the wording of any of the questions on this**

**form.**

**ICMJE DISCLOSURE FORM**

**Date:** January 8, 2025

**Your Name:** Benjamin Rabin

**Manuscript** Title: Global gram-negative diabetic foot infection prevalence and its associations with climate:
a systematic review and meta-analysis

**Manuscript number (if known):__________________________________________________________________**

**In the interest of transparency, we ask you to disclose all relationships/activities/interests listed below that are**

**related to the content of your manuscript. “Related” means any relation with for-profit or not-for-profit third**

**parties whose interests may be affected by the content of the manuscript. Disclosure represents a commitment**

**to transparency and does not necessarily indicate a bias. If you are in doubt about whether to list a relationship/activity/interest, it is preferable that you do so.**

**The following questions apply to the** **author’s relationships/activities/interests as they relate to the current**

**manuscript only.**

**The author’s relationships/activities/interests should be defined broadly. For example, if your manuscript pertains**

**to the epidemiology of hypertension, you should declare all relationships with manufacturers of antihypertensive medication, even if that medication is not mentioned in the manuscript.**

**In item #1 below, report all support for the work reported in this manuscript without time limit. For all other items,**

**the time frame for disclosure is the past 36 months.**

|  |  | **Name all entities with whom you have this relationship or indicate none (add rows as needed)** | | **Specifications/Comments**  **(e.g., if payments were made to you or to your institution)** |
| --- | --- | --- | --- | --- |
| **Time frame: Since the initial planning of the work** | | | | |
| 1 | All support for the present manuscript (e.g., funding, provision of study materials, medical writing, article processing charges, etc.)  **No time limit for this item.** | None | |  |
| **Time frame: past 36 months** | | | | |
| 2 | Grants or contracts from any entity (if not indicated in item #1 above). | None |  | |
| 3 | Royalties or licenses | None |  | |
| 4 | Consulting fees | None |  | |
| 5 | Payment or honoraria for lectures, presentations, speakers bureaus, manuscript writing or educational events | None |  | |
| 6 | Payment for expert testimony | None |  | |
| 7 | Support for attending meetings and/or travel | None |  | |
| 8 | Patents planned, issued or pending | None |  | |
| 9 | Participation on a Data  Safety Monitoring Board or Advisory Board | None |  | |
| 10 | Leadership or fiduciary role in other board, society, committee or advocacy group, paid or unpaid | None |  | |
| 11 | Stock or stock options | None |  | |
| 12 | Receipt of equipment, materials, drugs, medical writing, gifts or other services | None |  | |
| 13 | Other financial or non-financial interests | None |  | |

**Please place an “X” next to the following statement to indicate your agreement:**

**X	I certify that I have answered every question and have not altered the wording of any of the questions on this**

**form.**

**ICMJE DISCLOSURE FORM**

**Date:** 01/13/25**_______________________________________________________________________________**

**Your Name:** Ellen Martinson**____________________________________________________________________**

**Manuscript Title:** Global gram-negative diabetic foot infection prevalence and its associations with climate: a systematic review and meta-analysi**s______________________________________________________________________**

**Manuscript number (if known):__________________________________________________________________**

**In the interest of transparency, we ask you to disclose all relationships/activities/interests listed below that are**

**related to the content of your manuscript. “Related” means any relation with for-profit or not-for-profit third**

**parties whose interests may be affected by the content of the manuscript. Disclosure represents a commitment**

**to transparency and does not necessarily indicate a bias. If you are in doubt about whether to list a relationship/activity/interest, it is preferable that you do so.**

**The following questions apply to the author’s relationships/activities/interests as they relate to the current**

**manuscript only.**

**The author’s relationships/activities/interests should be defined broadly. For example, if your manuscript pertains**

**to the epidemiology of hypertension, you should declare all relationships with manufacturers of antihypertensive medication, even if that medication is not mentioned in the manuscript.**

**In item #1 below, report all support for the work reported in this manuscript without time limit. For all other items,**

**the time frame for disclosure is the past 36 months.**

|  |  | **Name all entities with whom you have this relationship or indicate none (add rows as needed)** | | **Specifications/Comments**  **(e.g., if payments were made to you or to your institution)** |
| --- | --- | --- | --- | --- |
| **Time frame: Since the initial planning of the work** | | | | |
| 1 | All support for the present manuscript (e.g., funding, provision of study materials, medical writing, article processing charges, etc.)  **No time limit for this item.** | __X__None | |  |
| **Time frame: past 36 months** | | | | |
| 2 | Grants or contracts from any entity (if not indicated in item #1 above). | __X__None |  | |
| 3 | Royalties or licenses | __X__None |  | |
| 4 | Consulting fees | __X__None |  | |
| 5 | Payment or honoraria for lectures, presentations, speakers bureaus, manuscript writing or educational events | __X__None |  | |
| 6 | Payment for expert testimony | __X__None |  | |
| 7 | Support for attending meetings and/or travel | __X__None |  | |
| 8 | Patents planned, issued or pending | __X__None |  | |
| 9 | Participation on a Data  Safety Monitoring Board or Advisory Board | __X__None |  | |
| 10 | Leadership or fiduciary role in other board, society, committee or advocacy group, paid or unpaid | __X__None |  | |
| 11 | Stock or stock options | __X__None |  | |
| 12 | Receipt of equipment, materials, drugs, medical writing, gifts or other services | __X__None |  | |
| 13 | Other financial or non-financial interests | __X__None |  | |

**Please place an “X” next to the following statement to indicate your agreement:**

**_X__ I certify that I have answered every question and have not altered the wording of any of the questions on this**

**form.**

**ICMJE DISCLOSURE FORM**

**Date:** January 8, 2025

**Your Name:** Mia S. White

**Manuscript** Title: Global gram-negative diabetic foot infection prevalence and its associations with climate:
a systematic review and meta-analysis

**Manuscript number (if known):__________________________________________________________________**

**In the interest of transparency, we ask you to disclose all relationships/activities/interests listed below that are**

**related to the content of your manuscript. “Related” means any relation with for-profit or not-for-profit third**

**parties whose interests may be affected by the content of the manuscript. Disclosure represents a commitment**

**to transparency and does not necessarily indicate a bias. If you are in doubt about whether to list a relationship/activity/interest, it is preferable that you do so.**

**The following questions apply to the author’s relationships/activities/interests as they relate to the current**

**manuscript only.**

**The author’s relationships/activities/interests should be defined broadly. For example, if your manuscript pertains**

**to the epidemiology of hypertension, you should declare all relationships with manufacturers of antihypertensive medication, even if that medication is not mentioned in the manuscript.**

**In item #1 below, report all support for the work reported in this manuscript without time limit. For all other items,**

**the time frame for disclosure is the past 36 months.**

|  |  | **Name all entities with whom you have this relationship or indicate none (add rows as needed)** | | **Specifications/Comments**  **(e.g., if payments were made to you or to your institution)** |
| --- | --- | --- | --- | --- |
| **Time frame: Since the initial planning of the work** | | | | |
| 1 | All support for the present manuscript (e.g., funding, provision of study materials, medical writing, article processing charges, etc.)  **No time limit for this item.** | None | |  |
| **Time frame: past 36 months** | | | | |
| 2 | Grants or contracts from any entity (if not indicated in item #1 above). | None |  | |
| 3 | Royalties or licenses | None |  | |
| 4 | Consulting fees | None |  | |
| 5 | Payment or honoraria for lectures, presentations, speakers bureaus, manuscript writing or educational events | None |  | |
| 6 | Payment for expert testimony | None |  | |
| 7 | Support for attending meetings and/or travel | None |  | |
| 8 | Patents planned, issued or pending | None |  | |
| 9 | Participation on a Data  Safety Monitoring Board or Advisory Board | None |  | |
| 10 | Leadership or fiduciary role in other board, society, committee or advocacy group, paid or unpaid | None |  | |
| 11 | Stock or stock options | None |  | |
| 12 | Receipt of equipment, materials, drugs, medical writing, gifts or other services | None |  | |
| 13 | Other financial or non-financial interests | None |  | |

**Please place an “X” next to the following statement to indicate your agreement:**

**X	I certify that I have answered every question and have not altered the wording of any of the questions on this**

**form.**

**ICMJE DISCLOSURE FORM**

**Date: 02/13/2024_______________________________________________________**

**Your Name:Baffour Otchere_______________________________________________________**

**Manuscript Title: Global gram-negative diabetic foot infection prevalence and its associations with climate: a systematic review and meta-analysis**

**Manuscript number (if known):__________________________________________________________________**

**In the interest of transparency, we ask you to disclose all relationships/activities/interests listed below that are**

**related to the content of your manuscript. “Related” means any relation with for-profit or not-for-profit third**

**parties whose interests may be affected by the content of the manuscript. Disclosure represents a commitment**

**to transparency and does not necessarily indicate a bias. If you are in doubt about whether to list a relationship/activity/interest, it is preferable that you do so.**

**The following questions apply to the author’s relationships/activities/interests as they relate to the current**

**manuscript only.**

**The author’s relationships/activities/interests should be defined broadly. For example, if your manuscript pertains**

**to the epidemiology of hypertension, you should declare all relationships with manufacturers of antihypertensive medication, even if that medication is not mentioned in the manuscript.**

**In item #1 below, report all support for the work reported in this manuscript without time limit. For all other items,**

**the time frame for disclosure is the past 36 months.**

|  |  | **Name all entities with whom you have this relationship or indicate none (add rows as needed)** | | **Specifications/Comments**  **(e.g., if payments were made to you or to your institution)** |
| --- | --- | --- | --- | --- |
| **Time frame: Since the initial planning of the work** | | | | |
| 1 | All support for the present manuscript (e.g., funding, provision of study materials, medical writing, article processing charges, etc.)  **No time limit for this item.** | ____None | |  |
| **Time frame: past 36 months** | | | | |
| 2 | Grants or contracts from any entity (if not indicated in item #1 above). | ____None |  | |
| 3 | Royalties or licenses | ____None |  | |
| 4 | Consulting fees | ____None |  | |
| 5 | Payment or honoraria for lectures, presentations, speakers bureaus, manuscript writing or educational events | ____None |  | |
| 6 | Payment for expert testimony | ____None |  | |
| 7 | Support for attending meetings and/or travel | ____None |  | |
| 8 | Patents planned, issued or pending | ____None |  | |
| 9 | Participation on a Data  Safety Monitoring Board or Advisory Board | ____None |  | |
| 10 | Leadership or fiduciary role in other board, society, committee or advocacy group, paid or unpaid | ____None |  | |
| 11 | Stock or stock options | ____None |  | |
| 12 | Receipt of equipment, materials, drugs, medical writing, gifts or other services | ____None |  | |
| 13 | Other financial or non-financial interests | ____None |  | |

**Please place an “X” next to the following statement to indicate your agreement:**

**X I certify that I have answered every question and have not altered the wording of any of the questions on this**

**form.**

**ICMJE DISCLOSURE FORM**

**Date:** 1/9/2025

**Your Name:** Julia Raymond

**Manuscript Title:** Global gram-negative diabetic foot infection prevalence and its associations with climate: a systematic review and meta-analysis

**Manuscript number (if known):__________________________________________________________________**

**In the interest of transparency, we ask you to disclose all relationships/activities/interests listed below that are**

**related to the content of your manuscript. “Related” means any relation with for-profit or not-for-profit third**

**parties whose interests may be affected by the content of the manuscript. Disclosure represents a commitment**

**to transparency and does not necessarily indicate a bias. If you are in doubt about whether to list a relationship/activity/interest, it is preferable that you do so.**

**The following questions apply to the author’s relationships/activities/interests as they relate to the current**

**manuscript only.**

**The author’s relationships/activities/interests should be defined broadly. For example, if your manuscript pertains**

**to the epidemiology of hypertension, you should declare all relationships with manufacturers of antihypertensive medication, even if that medication is not mentioned in the manuscript.**

**In item #1 below, report all support for the work reported in this manuscript without time limit. For all other items,**

**the time frame for disclosure is the past 36 months.**

|  |  | **Name all entities with whom you have this relationship or indicate none (add rows as needed)** | | **Specifications/Comments**  **(e.g., if payments were made to you or to your institution)** |
| --- | --- | --- | --- | --- |
| **Time frame: Since the initial planning of the work** | | | | |
| 1 | All support for the present manuscript (e.g., funding, provision of study materials, medical writing, article processing charges, etc.)  **No time limit for this item.** | ____None | |  |
| **Time frame: past 36 months** | | | | |
| 2 | Grants or contracts from any entity (if not indicated in item #1 above). | ____None |  | |
| 3 | Royalties or licenses | ____None |  | |
| 4 | Consulting fees | ____None |  | |
| 5 | Payment or honoraria for lectures, presentations, speakers bureaus, manuscript writing or educational events | ____None |  | |
| 6 | Payment for expert testimony | ____None |  | |
| 7 | Support for attending meetings and/or travel | ____None |  | |
| 8 | Patents planned, issued or pending | ____None |  | |
| 9 | Participation on a Data  Safety Monitoring Board or Advisory Board | ____None |  | |
| 10 | Leadership or fiduciary role in other board, society, committee or advocacy group, paid or unpaid | ____None |  | |
| 11 | Stock or stock options | ____None |  | |
| 12 | Receipt of equipment, materials, drugs, medical writing, gifts or other services | ____None |  | |
| 13 | Other financial or non-financial interests | ____None |  | |

**Please place an “X” next to the following statement to indicate your agreement:**

__X **I certify that I have answered every question and have not altered the wording of any of the questions on this**

**form.**

**ICMJE DISCLOSURE FORM**

**Date: 25JAN2025_____________________________________________________________________________**

**Your Name: Jillian Dunbar______________________________________________________________________**

**Manuscript Title: Global gram-negative diabetic foot infection prevalence and its associations with climate: a systematic review and meta-analysis_______________________________________________________________________**

**Manuscript number (if known):__________________________________________________________________**

**In the interest of transparency, we ask you to disclose all relationships/activities/interests listed below that are**

**related to the content of your manuscript. “Related” means any relation with for-profit or not-for-profit third**

**parties whose interests may be affected by the content of the manuscript. Disclosure represents a commitment**

**to transparency and does not necessarily indicate a bias.  If you are in doubt about whether to list a relationship/activity/interest, it is preferable that you do so.**

**The following questions apply to the author’s relationships/activities/interests as they relate to the current**

**manuscript only.**

**The author’s relationships/activities/interests should be defined broadly. For example, if your manuscript pertains**

**to the epidemiology of hypertension, you should declare all relationships with manufacturers of antihypertensive medication, even if that medication is not mentioned in the manuscript.**

**In item #1 below, report all support for the work reported in this manuscript without time limit.  For all other items,**

**the time frame for disclosure is the past 36 months.**

|  |  | **Name all entities with whom you have this relationship or indicate none (add rows as needed)** | | **Specifications/Comments**  **(e.g., if payments were made to you or to your institution)** |
| --- | --- | --- | --- | --- |
| **Time frame: Since the initial planning of the work** | | | | |
| 1 | All support for the present manuscript (e.g., funding, provision of study materials, medical writing, article processing charges, etc.)  **No time limit for this item.** | _X__None | |  |
| **Time frame: past 36 months** | | | | |
| 2 | Grants or contracts from any entity (if not indicated in item #1 above). | _X__None |  | |
| 3 | Royalties or licenses | _X__None |  | |
| 4 | Consulting fees | _X__None |  | |
| 5 | Payment or honoraria for lectures, presentations, speakers bureaus, manuscript writing or educational events | _X__None |  | |
| 6 | Payment for expert testimony | _X__None |  | |
| 7 | Support for attending meetings and/or travel | _X__None |  | |
| 8 | Patents planned, issued or pending | _X__None |  | |
| 9 | Participation on a Data  Safety Monitoring Board or Advisory Board | _X__None |  | |
| 10 | Leadership or fiduciary role in other board, society, committee or advocacy group, paid or unpaid | _X__None |  | |
| 11 | Stock or stock options | _X__None |  | |
| 12 | Receipt of equipment,   materials, drugs, medical writing, gifts or other services | _X__None |  | |
| 13 | Other financial or non-financial interests | ___None |  | |
|  |  | _X_ US Navy non-financial interest. | “The views expressed are those of the author(s) and do not reflect the official policy or position of the US Navy, Department of Defense or the US Government” | |

**Please place an “X” next to the following statement to indicate your agreement:**

**_X_  I certify that I have answered every question and have not altered the wording of any of the questions on this**

**Form.**

**ICMJE DISCLOSURE FORM**

**Date: January 7th, 2025**

**Your Name: Meg McAloon**

**Manuscript Title: Global gram-negative diabetic foot infection prevalence and its associations with**

**climate: a systematic review and meta-analysis**

**Manuscript number (if known):__________________________________________________________________**

**In the interest of transparency, we ask you to disclose all relationships/activities/interests listed below that are**

**related to the content of your manuscript. “Related” means any relation with for-profit or not-for-profit third**

**parties whose interests may be affected by the content of the manuscript. Disclosure represents a commitment**

**to transparency and does not necessarily indicate a bias.  If you are in doubt about whether to list a relationship/activity/interest, it is preferable that you do so.**

**The following questions apply to the author’s relationships/activities/interests as they relate to the current**

**manuscript only.**

**The author’s relationships/activities/interests should be defined broadly. For example, if your manuscript pertains**

**to the epidemiology of hypertension, you should declare all relationships with manufacturers of antihypertensive medication, even if that medication is not mentioned in the manuscript.**

**In item #1 below, report all support for the work reported in this manuscript without time limit.  For all other items,**

**the time frame for disclosure is the past 36 months.**

|  |  | **Name all entities with whom you have this relationship or indicate none (add rows as needed)** | | **Specifications/Comments**  **(e.g., if payments were made to you or to your institution)** |
| --- | --- | --- | --- | --- |
| **Time frame: Since the initial planning of the work** | | | | |
| 1 | All support for the present manuscript (e.g., funding, provision of study materials, medical writing, article processing charges, etc.)  **No time limit for this item.** | None | |  |
| **Time frame: past 36 months** | | | | |
| 2 | Grants or contracts from any entity (if not indicated in item #1 above). | None |  | |
| 3 | Royalties or licenses | None |  | |
| 4 | Consulting fees | None |  | |
| 5 | Payment or honoraria for lectures, presentations, speakers bureaus, manuscript writing or educational events | None |  | |
| 6 | Payment for expert testimony | None |  | |
| 7 | Support for attending meetings and/or travel | None |  | |
| 8 | Patents planned, issued or pending | None |  | |
| 9 | Participation on a Data  Safety Monitoring Board or Advisory Board | None |  | |
| 10 | Leadership or fiduciary role in other board, society, committee or advocacy group, paid or unpaid | None |  | |
| 11 | Stock or stock options | None |  | |
| 12 | Receipt of equipment,   materials, drugs, medical writing, gifts or other services | None |  | |
| 13 | Other financial or non-financial interests | None |  | |

**Please place an “X” next to the following statement to indicate your agreement:**

**X  I certify that I have answered every question and have not altered the wording of any of the questions on this**

**form.**

**ICMJE DISCLOSURE FORM**

**Date: 11^th^ February 2025**

**Your Name: Priyanka A Bhanushali**

**Manuscript Title: Global gram-negative diabetic foot infection prevalence and its associations with climate: a systematic review and meta-analysis**

**Manuscript number (if known):__________________________________________________________________**

**In the interest of transparency, we ask you to disclose all relationships/activities/interests listed below that are**

**related to the content of your manuscript. “Related” means any relation with for-profit or not-for-profit third**

**parties whose interests may be affected by the content of the manuscript. Disclosure represents a commitment**

**to transparency and does not necessarily indicate a bias.  If you are in doubt about whether to list a relationship/activity/interest, it is preferable that you do so.**

**The following questions apply to the author’s relationships/activities/interests as they relate to the current**

**manuscript only.**

**The author’s relationships/activities/interests should be defined broadly. For example, if your manuscript pertains**

**to the epidemiology of hypertension, you should declare all relationships with manufacturers of antihypertensive medication, even if that medication is not mentioned in the manuscript.**

**In item #1 below, report all support for the work reported in this manuscript without time limit.  For all other items,**

**the time frame for disclosure is the past 36 months.**

|  |  | **Name all entities with whom you have this relationship or indicate none (add rows as needed)** | | **Specifications/Comments**  **(e.g., if payments were made to you or to your institution)** |
| --- | --- | --- | --- | --- |
| **Time frame: Since the initial planning of the work** | | | | |
| 1 | All support for the present manuscript (e.g., funding, provision of study materials, medical writing, article processing charges, etc.)  **No time limit for this item.** | None | |  |
| **Time frame: past 36 months** | | | | |
| 2 | Grants or contracts from any entity (if not indicated in item #1 above). | None |  | |
| 3 | Royalties or licenses | None |  | |
| 4 | Consulting fees | None |  | |
| 5 | Payment or honoraria for lectures, presentations, speakers bureaus, manuscript writing or educational events | None |  | |
| 6 | Payment for expert testimony | None |  | |
| 7 | Support for attending meetings and/or travel | None |  | |
| 8 | Patents planned, issued or pending | None |  | |
| 9 | Participation on a Data  Safety Monitoring Board or Advisory Board | None |  | |
| 10 | Leadership or fiduciary role in other board, society, committee or advocacy group, paid or unpaid | None |  | |
| 11 | Stock or stock options | None |  | |
| 12 | Receipt of equipment,   materials, drugs, medical writing, gifts or other services | None |  | |
| 13 | Other financial or non-financial interests | None |  | |

**Please place an “X” next to the following statement to indicate your agreement:**

**_X_  I certify that I have answered every question and have not altered the wording of any of the questions on this**

**form.**

**ICMJE DISCLOSURE FORM**

**Date:** 1/18/2025

**Your Name:** Kyra Urquhart-Foster

**Manuscript Title:** Global gram-negative diabetic foot infection prevalence and its associations with climate: a systematic review and meta-analysis

**Manuscript number (if known):__________________________________________________________________**

**In the interest of transparency, we ask you to disclose all relationships/activities/interests listed below that are**

**related to the content of your manuscript. “Related” means any relation with for-profit or not-for-profit third**

**parties whose interests may be affected by the content of the manuscript. Disclosure represents a commitment**

**to transparency and does not necessarily indicate a bias.  If you are in doubt about whether to list a relationship/activity/interest, it is preferable that you do so.**

**The following questions apply to the author’s relationships/activities/interests as they relate to the current**

**manuscript only.**

**The author’s relationships/activities/interests should be defined broadly. For example, if your manuscript pertains**

**to the epidemiology of hypertension, you should declare all relationships with manufacturers of antihypertensive medication, even if that medication is not mentioned in the manuscript.**

**In item #1 below, report all support for the work reported in this manuscript without time limit.  For all other items,**

**the time frame for disclosure is the past 36 months.**

|  |  | **Name all entities with whom you have this relationship or indicate none (add rows as needed)** | | **Specifications/Comments**  **(e.g., if payments were made to you or to your institution)** |
| --- | --- | --- | --- | --- |
| **Time frame: Since the initial planning of the work** | | | | |
| 1 | All support for the present manuscript (e.g., funding, provision of study materials, medical writing, article processing charges, etc.)  **No time limit for this item.** | None | |  |
| **Time frame: past 36 months** | | | | |
| 2 | Grants or contracts from any entity (if not indicated in item #1 above). | None |  | |
| 3 | Royalties or licenses | None |  | |
| 4 | Consulting fees | None |  | |
| 5 | Payment or honoraria for lectures, presentations, speakers bureaus, manuscript writing or educational events | None |  | |
| 6 | Payment for expert testimony | None |  | |
| 7 | Support for attending meetings and/or travel | None |  | |
| 8 | Patents planned, issued or pending | None |  | |
| 9 | Participation on a Data  Safety Monitoring Board or Advisory Board | None |  | |
| 10 | Leadership or fiduciary role in other board, society, committee or advocacy group, paid or unpaid | None |  | |
| 11 | Stock or stock options | None |  | |
| 12 | Receipt of equipment,   materials, drugs, medical writing, gifts or other services | None |  | |
| 13 | Other financial or non-financial interests | None |  | |

**Please place an “X” next to the following statement to indicate your agreement:**

**X   I certify that I have answered every question and have not altered the wording of any of the questions on this**

**form.**

**ICMJE DISCLOSURE FORM**

**Date: January 14th, 2025**

**Your Name: Maya Fayfman**

**Manuscript Title: Global gram-negative diabetic foot infection prevalence and its associations with**

**climate: a systematic review and meta-analysis**

**Manuscript number (if known):__________________________________________________________________**

**In the interest of transparency, we ask you to disclose all relationships/activities/interests listed below that are**

**related to the content of your manuscript. “Related” means any relation with for-profit or not-for-profit third**

**parties whose interests may be affected by the content of the manuscript. Disclosure represents a commitment**

**to transparency and does not necessarily indicate a bias.  If you are in doubt about whether to list a relationship/activity/interest, it is preferable that you do so.**

**The following questions apply to the author’s relationships/activities/interests as they relate to the current**

**manuscript only.**

**The author’s relationships/activities/interests should be defined broadly. For example, if your manuscript pertains**

**to the epidemiology of hypertension, you should declare all relationships with manufacturers of antihypertensive medication, even if that medication is not mentioned in the manuscript.**

**In item #1 below, report all support for the work reported in this manuscript without time limit.  For all other items,**

**the time frame for disclosure is the past 36 months.**

|  |  | **Name all entities with whom you have this relationship or indicate none (add rows as needed)** | | **Specifications/Comments**  **(e.g., if payments were made to you or to your institution)** |
| --- | --- | --- | --- | --- |
| **Time frame: Since the initial planning of the work** | | | | |
| 1 | All support for the present manuscript (e.g., funding, provision of study materials, medical writing, article processing charges, etc.)  **No time limit for this item.** | None | |  |
| **Time frame: past 36 months** | | | | |
| 2 | Grants or contracts from any entity (if not indicated in item #1 above). | None |  | |
| 3 | Royalties or licenses | None |  | |
| 4 | Consulting fees | None |  | |
| 5 | Payment or honoraria for lectures, presentations, speakers bureaus, manuscript writing or educational events | None |  | |
| 6 | Payment for expert testimony | None |  | |
| 7 | Support for attending meetings and/or travel | None |  | |
| 8 | Patents planned, issued or pending | None |  | |
| 9 | Participation on a Data  Safety Monitoring Board or Advisory Board | None |  | |
| 10 | Leadership or fiduciary role in other board, society, committee or advocacy group, paid or unpaid | None |  | |
| 11 | Stock or stock options | None |  | |
| 12 | Receipt of equipment,   materials, drugs, medical writing, gifts or other services | None |  | |
| 13 | Other financial or non-financial interests | None |  | |

**Please place an “X” next to the following statement to indicate your agreement:**

**X  I certify that I have answered every question and have not altered the wording of any of the questions on this**

**form.**

| ICMJE DISCLOSURE FORM | |
| --- | --- |
| **Date:** | 2/14/2025 |
| **Your Name:** | Mohammed K Ali |
| **Manuscript Title:** | **Global gram-negative diabetic foot infection prevalence and its associations with climate: a systematic review and meta-analysis** |
| **Manuscript Number (if known):** | Click or tap here to enter text. |
| In the interest of transparency, we ask you to disclose all relationships/activities/interests listed below that are related to the content of your manuscript. “Related” means any relation with for-profit or not-for-profit third parties whose interests may be affected by the content of the manuscript. Disclosure represents a commitment to transparency and does not necessarily indicate a bias. If you are in doubt about whether to list a relationship/activity/interest, it is preferable that you do so.  The author’s relationships/activities/interests should be defined broadly. For example, if your manuscript pertains to the epidemiology of hypertension, you should declare all relationships with manufacturers of antihypertensive medication, even if that medication is not mentioned in the manuscript.  In item #1 below, report all support for the work reported in this manuscript without time limit. For all other items, the time frame for disclosure is the past 36 months. | |

|  | | | **Name all entities with whom you have this relationship or indicate none (add rows as needed)** | **Specifications/Comments (e.g., if payments were made to you or to your institution)** |
| --- | --- | --- | --- | --- |
| **Time frame: Since the initial planning of the work** | | | | |
| **1** | All support for the present manuscript (e.g., funding, provision of study materials, medical writing, article processing charges, etc.)  **No time limit for this item.** | | ☒ \| **None** \| Click the tab key to add additional rows. | |
| **Time frame: past 36 months** | | | | |
| **2** | | Grants or contracts from any entity (if not indicated in item #1 above). | ☒ \| **None** | |
| **3** | | Royalties or licenses | ☒ \| **None** | |
| **4** | | Consulting fees | ☒ \| **None** | |
| **5** | | Payment or honoraria for lectures, presentations, speakers bureaus, manuscript writing or educational events | ☒ \| **None** | |
| **6** | | Payment for expert testimony | ☒ \| **None** | |
| **7** | | Support for attending meetings and/or travel | ☒ \| **None** | |
| **8** | | Patents planned, issued or pending | ☒ \| **None** | |
| **9** | | Participation on a Data Safety Monitoring Board or Advisory Board | ☒ \| **None** | |
| **10** | | Leadership or fiduciary role in other board, society, committee or advocacy group, paid or unpaid | ☒ \| **None** | |
| **11** | | Stock or stock options | ☒ \| **None** | |
| **12** | | Receipt of equipment, materials, drugs, medical writing, gifts or other services | ☒ \| **None** | |
| **13** | | Other financial or non-financial interests | ☒ \| **None** | |
| **Please place an “X” next to the following statement to indicate your agreement:** | | | | |
| ☒ | | I certify that I have answered every question and have not altered the wording of any of the questions on this form. | | |

**ICMJE DISCLOSURE FORM**

**Date:__09 JANUARY 2025________________________________________________________________________**

**Your Name: ERIC SENNEVILLE____________________________________________**

**Manuscript Title:** Global gram-negative diabetic foot infection prevalence and its associations with climate: a systematic review and meta-analysis

**Manuscript number (if known):__________________________________________________________________**

**In the interest of transparency, we ask you to disclose all relationships/activities/interests listed below that are**

**related to the content of your manuscript. “Related” means any relation with for-profit or not-for-profit third**

**parties whose interests may be affected by the content of the manuscript. Disclosure represents a commitment**

**to transparency and does not necessarily indicate a bias. If you are in doubt about whether to list a relationship/activity/interest, it is preferable that you do so.**

**The following questions apply to the author’s relationships/activities/interests as they relate to the current**

**manuscript only.**

**The author’s relationships/activities/interests should be defined broadly. For example, if your manuscript pertains**

**to the epidemiology of hypertension, you should declare all relationships with manufacturers of antihypertensive medication, even if that medication is not mentioned in the manuscript.**

**In item #1 below, report all support for the work reported in this manuscript without time limit. For all other items,**

**the time frame for disclosure is the past 36 months.**

|  |  | **Name all entities with whom you have this relationship or indicate none (add rows as needed)** | | **Specifications/Comments**  **(e.g., if payments were made to you or to your institution)** |
| --- | --- | --- | --- | --- |
| **Time frame: Since the initial planning of the work** | | | | |
| 1 | All support for the present manuscript (e.g., funding, provision of study materials, medical writing, article processing charges, etc.)  **No time limit for this item.** | ____None | |  |
| **Time frame: past 36 months** | | | | |
| 2 | Grants or contracts from any entity (if not indicated in item #1 above). | ____None |  | |
| 3 | Royalties or licenses | ____None |  | |
| 4 | Consulting fees | ____None |  | |
| 5 | Payment or honoraria for lectures, presentations, speakers bureaus, manuscript writing or educational events | ____None |  | |
| 6 | Payment for expert testimony | ____None |  | |
| 7 | Support for attending meetings and/or travel | ____None |  | |
| 8 | Patents planned, issued or pending | ____None |  | |
| 9 | Participation on a Data  Safety Monitoring Board or Advisory Board | ____None |  | |
| 10 | Leadership or fiduciary role in other board, society, committee or advocacy group, paid or unpaid | CHAIR OF THE IWGDF/IFDSA GUIDELINES FOR THE DIAGNOSIS AND MANAGEMENT OF DIABETIC FOOT INFECTION (2024 EDITION) |  | |
| 11 | Stock or stock options | ____None |  | |
| 12 | Receipt of equipment, materials, drugs, medical writing, gifts or other services | ____None |  | |
| 13 | Other financial or non-financial interests | ____None |  | |

**Please place an “X” next to the following statement to indicate your agreement:**

**X I certify that I have answered every question and have not altered the wording of any of the questions on this**

**form.**

**ICMJE DISCLOSURE FORM**

**Date: January 7th, 2025**

**Your Name: Rodrigo M Carrillo-Larco**

**Manuscript Title: Global gram-negative diabetic foot infection prevalence and its associations with**

**climate: a systematic review and meta-analysis**

**Manuscript number (if known):__________________________________________________________________**

**In the interest of transparency, we ask you to disclose all relationships/activities/interests listed below that are**

**related to the content of your manuscript. “Related” means any relation with for-profit or not-for-profit third**

**parties whose interests may be affected by the content of the manuscript. Disclosure represents a commitment**

**to transparency and does not necessarily indicate a bias.  If you are in doubt about whether to list a relationship/activity/interest, it is preferable that you do so.**

**The following questions apply to the author’s relationships/activities/interests as they relate to the current**

**manuscript only.**

**The author’s relationships/activities/interests should be defined broadly. For example, if your manuscript pertains**

**to the epidemiology of hypertension, you should declare all relationships with manufacturers of antihypertensive medication, even if that medication is not mentioned in the manuscript.**

**In item #1 below, report all support for the work reported in this manuscript without time limit.  For all other items,**

**the time frame for disclosure is the past 36 months.**

|  |  | **Name all entities with whom you have this relationship or indicate none (add rows as needed)** | | **Specifications/Comments**  **(e.g., if payments were made to you or to your institution)** |
| --- | --- | --- | --- | --- |
| **Time frame: Since the initial planning of the work** | | | | |
| 1 | All support for the present manuscript (e.g., funding, provision of study materials, medical writing, article processing charges, etc.)  **No time limit for this item.** | None | |  |
| **Time frame: past 36 months** | | | | |
| 2 | Grants or contracts from any entity (if not indicated in item #1 above). | None |  | |
| 3 | Royalties or licenses | None |  | |
| 4 | Consulting fees | None |  | |
| 5 | Payment or honoraria for lectures, presentations, speakers bureaus, manuscript writing or educational events | None |  | |
| 6 | Payment for expert testimony | None |  | |
| 7 | Support for attending meetings and/or travel | None |  | |
| 8 | Patents planned, issued or pending | None |  | |
| 9 | Participation on a Data  Safety Monitoring Board or Advisory Board | None |  | |
| 10 | Leadership or fiduciary role in other board, society, committee or advocacy group, paid or unpaid | None |  | |
| 11 | Stock or stock options | None |  | |
| 12 | Receipt of equipment,   materials, drugs, medical writing, gifts or other services | None |  | |
| 13 | Other financial or non-financial interests | None |  | |

**Please place an “X” next to the following statement to indicate your agreement:**

**X  I certify that I have answered every question and have not altered the wording of any of the questions on this**

**form.**
