## Supplementary material for "Global gram-negative diabetic foot infection prevalence and its associations with climate: a systematic review and meta-analysis": Prospero registration

### A systematic review and meta-analysis of the association between climate and gram-negative diabetic foot infections

To enable PROSPERO to focus on COVID-19 submissions, this registration record has undergone basic automated checks for eligibility and is published exactly as submitted. PROSPERO has never provided peer review, and usual checking by the PROSPERO team does not endorse content. Therefore, automatically published records should be treated as any other PROSPERO registration. Further detail is provided here.

#### Citation

Marcos Schechter, Mia White, Ben Rabin, Ellen Martinson, Priyanka Bhanushali, J Raymond, Jilian Dunbar, Baffour Otchere, Sophie Lockwood, Meg McAloon, Kyra Urquhart-Foster, Morgan Schafer. A systematic review and meta-analysis of the association between climate and gram-negative diabetic foot infections. PROSPERO 2022 CRD42022279019 Available from: https://www.crd.york.ac.uk/prospero/display_record.php?ID=CRD42022279019

#### Review question

P: patients with diabetic foot infection (soft tissue and/or osteomyelitis) who had a bacterial microbiology test (from soft tissue and/or bone collected at bedside and/or operating room).

I: any microbiology test including conventional and non-conventional methods (e.g., 16S)

C: climate patient lives in (temperate vs tropical/subtropical), using the healthcare facility as proxy. We will also use the Kopper-Geiger system to define climate.

O: proportion with gram-negative bacteria (main outcome) or Pseudomonas aeruginosa (secondary outcome) isolated

#### Searches

Timespam2010-2020

PubMed:

1.("Diabetic Foot"[MeSH Terms] AND ("infections"[MAJR] OR "soft tissue infect*"[Text Word] OR "Osteomyelitis"[Text Word] OR ulcer*[Text Word])) OR "diabetic foot infect*"[Text Word]

2."Foot"[MeSH Terms] AND ("Diabetes Complications"[MAJR] OR “diabetes mellitus/complications”[MAJR])

3. (("diabetic foot"[Text Word] OR (“diabetes”[TW] AND “foot”[TW])) AND (infect[Text Word] OR infected[TW] OR infection[TW] OR infections[TW] OR infecting[TW] OR ulcer*[TW] OR osteomyelitis[TW] OR “soft tissue infect*”[TW])) OR DFI[TW] OR DFU[TW]

4.#1OR#2OR#3

5."Bacterial Typing Techniques"[MeSH Terms] OR "bacterial typing"[Text Word] OR "bacterial identif*"[Text Word] OR "microbiology"[Text Word] OR "microbiology"[MeSH Subheading] OR "diagnosis"[Text Word] OR "diagnosis"[Subheading:NoExp] OR "isolation and purification"[MeSH Subheading] OR pathogen*[Text Word] OR "culture"[Text Word] OR (bacteria*[TW] AND classification[TW])

6.((#4 AND #5) AND (“Journal article”[PTYP] OR “journal article”[TW]) AND "2010/01/01"[PDat] : "3000/12/31"[PDat]) NOT (“PubMed books”[Filter]OR“editorial”[PTYP]OR“personal narrative”[PTYP])

Embase

1.‘Diabetic Foot’/exp AND ('infection'/exp/mj OR ‘soft tissue infection*’:ti, ab, kw OR ulcer*:ti, ab, kw)

2.'diabetic foot osteomyelitis'/exp

3.(‘diabetic foot*’:ti, ab, kw)

4.(‘diabetic foot*’ NEAR/3 (infect* OR ulcer* OR osteomyelitis)) OR ((diabetes NEAR/3 foot) AND (infect* OR ulcer* OR osteomyelitis):ti, ab, kw) OR DFI:ti, ab, kw OR DFU:ti, ab, kw

5.#1OR#2OR#3OR#4

6. 'bacterium identification'/exp OR bacteria*:ti, ab, kw OR (bacteria* NEAR/3 (identif* OR infect* OR classification OR diagnose OR diagnosis)) OR microbiology:ti, ab, kw OR 'microbiology'/exp/mj OR 'diagnosis'/exp/mj OR diagnosis:ti, ab, kw OR 'isolation and purification'/exp OR pathogen*:ti, ab, kw OR (identif* NEAR/3 pathogen*) OR Culture:ti, ab, kw

7.#5AND#6

#8 #7 AND (2010:pyOR 2011:pyOR2012:pyOR2013:pyOR 2014:pyOR 2015:py OR 2016:py OR 2017:py OR 2018:py OR 2019:py OR 2020:py) AND ([article]/lim OR [article in press]/lim OR [letter]/lim OR [review]/lim) NOT (([animals]/lim OR ‘animals’/exp OR animal*:ti, ab, kw)NOT([humans]/lim OR 'human'/exp OR human*:ti, ab, kw))

WebofScience:

1.(TS=("diabetic foot" NEAR/3 (infect* OR ulcer* OR osteomyelitis)) OR TS=((diabetes AND foot* NEAR/3 (infect* OR ulcer* OR osteomyelitis)))

2.TS=(bacteri*ORmicrobio*ORculture*OR pathogen*NEAR/3 (diagnosisORdiagnoseOR identi*OR classifOR etiology) )

3.#1AND#2

4.(#3)ANDDOCUMENT TYPES: (Article OR Early Access OR Letter OR Review)

Indexes=SCI-EXPANDED, SSCI, A&HCI, CPCI-S, CPCI-SSH, BKCI-S, BKCI-SSH, ESCI, CCR-EXPANDED, IC

AIM, IMSEAR, IMEMR, LILACS, WPRIM

(tw:(diabetes foot infection))OR(tw:(diabetic foot* bacteri*))OR(tw:(diabetic foot* microbiolog*))OR(tw:(diabetic foot* pathogen*))

#### Types of study to be included

There are no restrictions to study design. We will exclude reports the include <5 patients.

#### Condition or domain being studied

Diabetic foot infections

#### Participants/population

Patients with diabetic foot infection (soft tissue and/or osteomyelitis) who had a bacterial microbiology test (from soft tissue and/or bone collected at bedside and/or operating room).

#### Intervention(s), exposure(s)

Patients who had any sample from an infected diabetic foot sent for microbiological tests (including conventional and non-conventional methods) will be included. Patients that did not have a sample for microbiological tests will be excluded. Reports that include the proportion of patients with at least one gram-negative bacteria isolated will be included. Reports that only include patients with a specific type of bacteria will be excluded.

#### Comparator(s)/control

Climate patient lives in (temperate vs tropical/subtropical). The healthcare facility location will be used as proxy to the where the patients lives. We will also use the Kopper-Geiger system to define climate.

#### Main outcome(s)

Proportion of patients with at least one gram-negative bacteria isolated.

##### Measures of effect

We will calculate pooled proportions stratified by climate (temperate vs tropical/sub-tropical and Kopper-Geiger system definitions). Analyses will be conducted with the meta package for R. The pooled proportions will be calculated using a random intercept logistic regression model via the metaprop function. We will report fixed and random-effects model. Forest plots with 95% confidence intervals (CIs) will be built. The $I^2$ statistic was used to assess between study heterogeneity with P values based on the Q-statistic.

#### Additional outcome(s)

Our secondary outcomes will be the proportion of patients with Pseudomonas aeruginosa and Acinetobacter baumannii isolated.

##### Measures of effect

The measures of effect will be the same as the main outcome.

#### Data extraction (selection and coding)

We will include studies that report >5 unique patients with diabetic foot infection (soft tissue and/or osteomyelitis) who had a bacterial microbiology test (from soft tissue and/or bone collected at bedside and/or operating room). We will extract demographics (age, gender), mean/median A1c, prevalence of PAD, antibiotic exposure, infection severity classification, sample type (soft tissue, bone), in addition to microbiology results.

#### Risk of bias (quality) assessment

We will abstract data on (1) study design (2) patient selection [consecutive vs non-consecutive patients] (3) if PAD and antibiotic exposure are reported and (4) if infection severity is reported

#### Strategy for data synthesis

Analyses will be conducted with the meta package for R. The pooled proportions will be calculated using a random intercept logistic regression model via the metaprop function. We will report fixed and random-effects model. Forest plots with 95% confidence intervals (CIs) will be built. The $I^2$ statistic was used to assess between study heterogeneity with P values based on the Q-statistic.

#### Analysis of subgroups or subsets

Subset analyses for the main and secondary outcomes will be performed stratifying by the following characteristics: (1) prior antibiotic use (2) presence/absence of peripheral artery disease (3) specimen type - soft tissue vs bone, surgical vs non surgical (4) severity of infection

#### Contact details for further information

Marcos Schechter

#### Organisational affiliation of the review

Emory University

https://www.med.emory.edu/

#### Review team members and their organisational affiliations

Dr Marcos Schechter. Emory University

Mia White. Emory University

Ben Rabin. Emory University

Ellen Martinson. Emory University

Priyanka Bhanushali. Emory University

J Raymond. Emory University

Jilian Dunbar. Emory University

Baffour Otchere. Emory University

Sophie Lockwood. Emory University

Meg McAloon. Emory University

Kyra Urquhart-Foster. Augusta Medical College

Morgan Schafer. Emory University

#### Type and method of review

Epidemiologic, Systematic review

#### Anticipated or actual start date

01 March 2021

#### Anticipated completion date

01 December 2022

#### Funding sources/sponsors

None

Grant number(s)

State the funder, grant or award number and the date of award

Not applicable

#### Conflicts of interest

#### Language

English

#### Country

United States of America

#### Stage of review

Review Ongoing

#### Subject index terms status

Subject indexing assigned by CRD

#### Subject index terms

Anti-Bacterial Agents; Diabetes Mellitus; Diabetic Foot; Humans; Risk Factors

#### Date of registration in PROSPERO

27 June 2022

#### Date of first submission

16 June 2022

#### Stage of review at time of this submission

| **Stage** | **Started** | **Completed** |
|---|---|---|
| Preliminary searches | Yes | Yes |
| Piloting of the study selection process | No | No |
| Formal screening of search results against eligibility criteria | No | No |
| Data extraction | No | No |
| Risk of bias (quality) assessment | No | No |
| Data analysis | No | No |

*The record owner confirms that the information they have supplied for this submission is accurate and complete and they understand that deliberate provision of inaccurate information or omission of data may be construed as scientific misconduct.*

*The record owner confirms that they will update the status of the review when it is completed and will add publication details in due course.*

#### Versions

27 June 2022

27 June 2022
